# Single-nucleus multiomic landscape of congenital heart diseases reveals disease-specific genotypic profiles

**DOI:** 10.1101/2025.08.01.25332780

**Authors:** Dominika Lukovic, Mariann Gyöngyösi, Benjamen White, Emilie Han, Ena Hasimbegovic, Katrin Müller-Zlabinger, Artur Gynter, Veronika Mancikova, Imre J Pavo, Mathias Michelitsch, Ina Michel-Behnke

## Abstract

Tetralogy of Fallot (TOF) is the most common cyanotic congenital heart defect (CHD), whereas hypoplastic left heart syndrome (HLHS) represents 2-3% of all CHDs. We analyzed the single-nucleus multiome profile of right ventricle samples obtained from children during routine cardiac surgery for correction of TOF or staged surgical palliation for HLHS to define cell types and characterize gene regulation states in different cell clusters. Data were integrated with pre-existing controls and analyzed using Scanpy to identify clusters and annotated using automated tool and manual curation, pycisTopic to identify cell stats and cis-regulatory topics, and SCENIC+ for enhancer-driven gene regulatory network (eRegulon). Integrated RNA-seq analysis identified 22 different cell subtypes, including five cardiomyocyte phenotypes. TOF samples showed involvement in pathway networks of the cell cycle, DNA repair, DNA replication, and RNA metabolism, whereas gene expression in HLHS samples was related to extracellular matrix organization, anatomical structure development, cell adhesion, actin-myosin filament sliding, and contractile muscle fiber pathways. In addition, the gene expression fingerprints of endothelial, fibroblast, pericyte, immune, and neuronal cell nuclei from TOF and HLHS samples exhibited nuclei-specific significant de-regulation compared to controls. We found considerable heterogeneity among the transcriptomes of TOF and HLHS, explaining the diverse clinical phenotypes. These findings can enable the development of new gene-based interventions for specific CHDs.

---

Congenital heart defects (CHDs) are the most prevalent birth defect with only 15-20% of the genetic background being known. ^1,2^ Tetralogy of Fallot (TOF) is the most common cyanotic CHD, with a global incidence of 0.02–0.04%. ^3^ TOF is characterized by right ventricular outflow tract obstruction with valvular and/or subvalvular pulmonary stenosis; ventricular septal defect in which the aorta overrides the intraventricular septum, allowing venous blood to flow from the right ventricle to the systemic circulation; and right ventricular hypertrophy. Hypoplastic left heart syndrome (HLHS) represents 2-3% of all CHDs, with the typical feature of an underdeveloped left side of the heart (e.g., hypoplastic left ventricle), narrowed aorta, and malformation of the aortic and mitral valves, making it unable to provide the necessary amount of oxygenated blood to the main circulation. ^4^ Both TOF and HLHS are multigenic and phenotypically heterogeneous, and they could be associated with chromosomal aberrations or other heart defects, or paired with diverse clinical appearances, such as mental deterioration, facial dysmorphism, microcephaly, or cancer at early age, leading to multiple clinical phenotypes. ^5,6,7,8,9^ The pathophysiological mechanisms of the variable genetic backgrounds of these CHDs are poorly understood. Single-nucleus multiome RNA sequencing (RNA-seq) enables us to explore the transcriptional characteristics of pathophysiological conditions at the single-cell level. We aimed to characterize the genotypic variability of cardiac cells by analyzing the RNA profiles of nuclei of right ventricular biopsy samples from children with TOF and HLHS, and to define cell types and gene regulation states, including activation and maturation, in different cell clusters.

## Methods

### Data collection

In this single-center cohort study, we included pediatric patients undergoing clinically indicated cardiac surgery due to their CHD. Informed consent was obtained from the patient’s representative relatives because all patients were <8 years old. Tissue was only removed as required for the indicated surgery and only included in the biobank if the collected tissue was not required for clinically relevant diagnostic procedures. The study was approved by the institutional ethics committee of the Medical University of Vienna (EK. Nr. 1565/2019). Right ventricular samples were prospectively gathered from six patients with TOF and three patients with HLHS and stored either immediately in liquid nitrogen for isolation of RNA or in a 5% formaldehyde solution.

### Library construction and sequencing

Snap-frozen tissue samples were processed to generate single-nucleus ATAC and gene expression libraries following customized protocols from 10X Genomics. Nuclei isolation was performed using the Chromium Nuclei Isolation Kit with RNase Inhibitor (10X Genomics, PN-1000494) according to the user guide (CG000505, Chromium Nuclei Isolation Kit, UG, RevA), with slight modifications. Briefly, 200 µl of lysis buffer was added to the sample and dissociated using pestles provided by the vendor. After adding an additional 300 µl of lysis buffer, the tissue was homogenized by pipette-mixing and incubated on ice for 10 min. Next, the suspension was loaded onto nuclei isolation columns and centrifuged for 20 s at 16,000 rcf and 4°C. The flow-through was vortexed for 10 s and centrifuged for 3 min at 500 rcf and 4°C in a swinging bucket rotor. The supernatant was removed and the pellet gently resuspended in 500 µl of the supplied Debris Removal Buffer before centrifuging for 10 min at 700 rcf and 4°C. After removal of the supernatant, the pellet was washed once with 1 ml of the provided wash buffer, followed by centrifugation for 5 min at 500 rcf and 4°C. For multiplexing, we applied a customized approach based on barcoding with cholesterol-modified oligos (CMOs). Eight samples were pooled per single-nucleus sequencing reaction. For this, the nuclei were incubated after the first wash with unique CMO anchor-barcode and co-anchor and then washed twice with 1 ml of wash buffer by centrifugation for 5 min at 500 rcf and 4°C. The final nucleus pellet was taken up in 30 µl of Diluted Nuclei Buffer (10X Genomics) and strained once through a 30-µm pre-separation filter (Miltenyi, 130-041-407). Nuclei were counted using acridine orange/propidium iodide stain (Logos, F23001) and an automated dual fluorescence cell counter (Logos, LUNA-FL™). Equal numbers of nuclei were pooled per multiplexed reaction, resulting in a total of 16,000 nuclei per individual Chromium Next GEM Single Cell Multiome ATAC + Gene Expression (10X Genomics, PN-1000285) reaction. The pooled nuclei were transposed, loaded on a Chromium Next GEM Chip J (10X Genomics, PN-1000230), and run on the Chromium iX instrument as instructed by the manufacturer. ATAC and gene expression libraries were generated according to the user guide (CG000338, Chromium Next GEM Multiome ATAC GEX User Guide RevG).

The CMO hash library was generated by adding an additive primer to the cDNA amplification, subsequently purifying hash-DNA from the supernatant during cDNA cleanup, and ultimately amplifying CMO hash libraries with unique combinations of i5 and i7 indices. Library quality and size were assessed using the High Sensitivity DNA Analysis Kit (Agilent Technologies #5067-4626) and the Bioanalyzer 2100. For sequencing, gene expression libraries were pooled with 17% corresponding CMO hash libraries and sequenced on an Illumina NovaSeq6000 system using the following parameters: read 1, 28 cycles; i7, 10 cycles; i5, 10 cycles; read 2, 90 cycles. The ATAC libraries were sequenced on an Illumina NovaSeq6000 system using the following parameters: read 1N, 50 cycles; i7, 8 cycles; i5, 24 cycles; read 2N, 49 cycles.

### RNA-seq and snATAC-seq analysis

Single-nucleus ATAC sequencing (snATAC-seq) data were filtered for the presence of mitochondria (<25) and cells with a low number of genes by count (<6,800) using Scanpy. Cells were manually annotated according to the RNA-seq data and SCENIC+ package. PycisTopic was used for peak calling and to identify cell states and cis-regulatory topics. Consensus peaks were further filtered using pycisTopic (Log_unique_nr_frag min. 3.3, FRIP min. 0.45, and TSS_enrichment min. 5) and according to ENCODE blacklisted regions.^10^ Following this, SCENIC+ was used for enhancer-driven gene regulatory network (eRegulon) construction of the integrated snATAC-seq and RNA-seq data.^11^

Raw sequencing data were demultiplexed using bcl2fastq v2.20.0.422 (https://emea.support.illumina.com/sequencing/sequencing_software/bcl2fastq-conversion-software.html). The data were further processed in the cellranger count pipeline (version 7.1.0, 10X Genomics, Pleasanton, CA, USA).^12^ The GRCh38 assembly was used to align the sequenced reads to the human genome. Each cell barcode was assigned a donor identity based on further demultiplexing of hashtag barcodes. The demultiplexing was performed using a Gaussian mixture model fitted to hashtag barcode counts, inferring the probability that a given hashtag barcode read count is derived from background or constitutes a positive signal. ^13^ The processed RNA-seq data were integrated with pre-existing controls and analyzed using Scanpy, ^14^ to cluster and manually annotate cells (Table 1). Each cell cluster (main and cell subtypes) contained the pooled nuclei of the multiple biopsy samples for the same disease entity.

**Table 1.**
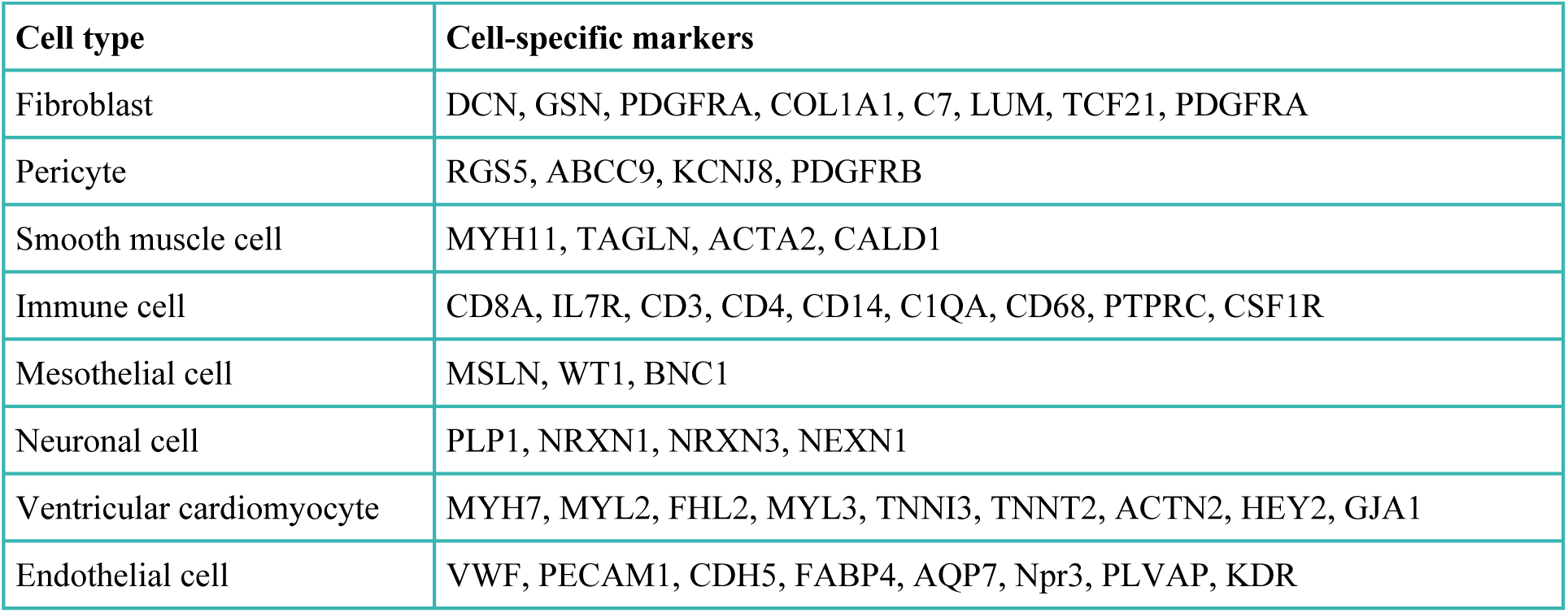
Cell type markers used for manual annotation

The publicly available interactive program for phenotyping cardiac cells was used (www.heartcellatlas.org)^15^, which describes five different cardiomyocytes and their main features (Suppl. Table S1).

The marker genes for subtypes of endothelial cells and fibroblasts were retrieved from CellTypist, Healthy_Adult_Heart.^16^

Due to the small numbers of vessels in the HLHS and TOF samples and the smooth muscle cell (SMC) expression overlapping with endothelial cells of arterial vessel origin (EC-arterial), SMCs were not analyzed for differences between CHD and controls (Suppl. Fig. S1).

Original data are submitted to GEO repository ([geo] GEO Submission (GSE300526) [NCBI tracking system #25232329].

### Comparing gene expression using Loupe Browser

Loupe Browser (version 8.1.2) is a freely downloadable program that was used to compare gene expression in the two CHD and control groups. The topological structure of the individual cell types, as well as singular or combined gene sets, were displayed by Uniform Manifold Approximation and Projection for Dimension Reduction (UMAP). The differences in gene expression between the groups were displayed in heat maps and violin plots. All significantly up- or down-regulated genes were included if the expression was common in each relevant cell phenotype. Heterogeneity of gene expression between the nuclei was determined if the genetic information of nuclei belonging to the same cell cluster was different. Gene selection was made based on the highest p-value. The p-values were adjusted by Benjamini-Hochberg correction for multiple testing.

We used gProfiler software (version 92, ^17,18^) to display the functional profiles of the significantly de-regulated genes. Reactome Pathway browser (reactome.org/PathwayBrowser) ^19,20^ displayed the roles and pathway connections of the significantly de-regulated genes in the HLHS and TOF groups compared to controls. In further downstream analysis, the protein-protein interaction network with functional enrichment analysis was created (string-db.org, version 12.0 ^21,22^) using the following enrichment display settings: maximum false discovery rate (FDR) ≤ 0.05, minimum signal ≥ 0.01, minimum strength ≥0.01, minimum count in network 2.

### Histology

Formalin-fixed, paraffin-embedded myocardial tissue sections were cut into 3-µm-thick slices and mounted on adhesive glass slides (X-tra® Adhesive Slides, Leica Biosystems). To assess cardiomyocyte morphology and cardiac fibrosis, standard staining with hematoxylin and eosin and picrosirius red (PSR) was performed according to established protocols. All stained slides were scanned in their entirety using a digital research slide scanner (Slideview VS200, Olympus, Tokyo, Japan). Quantitative analyses were performed using HALO (version 3.3.2541.184, Indica Labs, Albuquerque, New Mexico USA), QuPath ^23^, and OlyVIA (Olympus, Tokio, Japan) software. To assess cell area, 30 high-magnification images were taken of each scan. Afterwards, individual cells were manually traced on each image and the respective cell diameter and area recorded. To assess the proportionate fibrotic area from the PSR-stained samples, an annotation layer avoiding visible vessels was created. The proportion of fibrotic area for each sample was calculated as the PSR-positive percentage of the total assessed area.

## Results

### Clinical data and histology of the right ventricle

Six patients with TOF and three patients with HLHS were included in this study (Table 2). Right ventricular myocardial samples were obtained during routine cardiac surgery to correct TOF and staged surgical palliation for HLHS. Hematoxylin and eosin and picrosirius staining revealed a lower density of cell nuclei and higher proportion of fibrotic tissue in the HLHS samples (Fig. 1).

**Table 2.** Clinical characteristics and histological findings

|  | HLHS | TOF |
| --- | --- | --- |
| Clinical characteristics | n=3 | n=6 |
| Age, months | 18±13 | 12±12 |
| Sex, male | 2 (66.6%) | 4 (66.6%) |
| Height, cm | 75.5±12.4 | 69.1±9.0 |
| Weight, kg | 9.0±2.6 | 7.9±2.9 |
| BMI, kg/m<sup>2</sup> | 15.6±1.4 | 17.8±2.3 |
| BSA, m<sup>2</sup> | 0.45±0.08 | 0.39±0.09 |
| Median NT-proBNP, pg/ml (interquartile range) | 2512 (1111;5453) | 829 (565;829) |
| Echocardiography |  |  |
| Left ventricular end-diastolic volume, ml | 21±13 | 22±18 |
| Left ventricular end-systolic volume, ml | 6.1±4.1 | 6.3±3.2 |
| Left ventricular ejection fraction, % | 66.4±15.7 | 59.3±16.3 |
| Estimated maximal pulmonary arterial pressure, mmHg | 34.8±17.9 | 54.5±24.5 |
| **Histology** |  |  |
| Myocyte cell area, $\mu\text{m}^2$ | 126±18 | 139±72 |
| Myocardial fibrosis, % | 13.0±2.8 | 5.2±1.5 |

Values are given as mean±standard deviation or n (%) unless otherwise noted. BMI, body mass index; BSA, body surface area; HLHS, hypoplastic left heart syndrome; TOF, tetralogy of Fallot.

**Figure 1.**
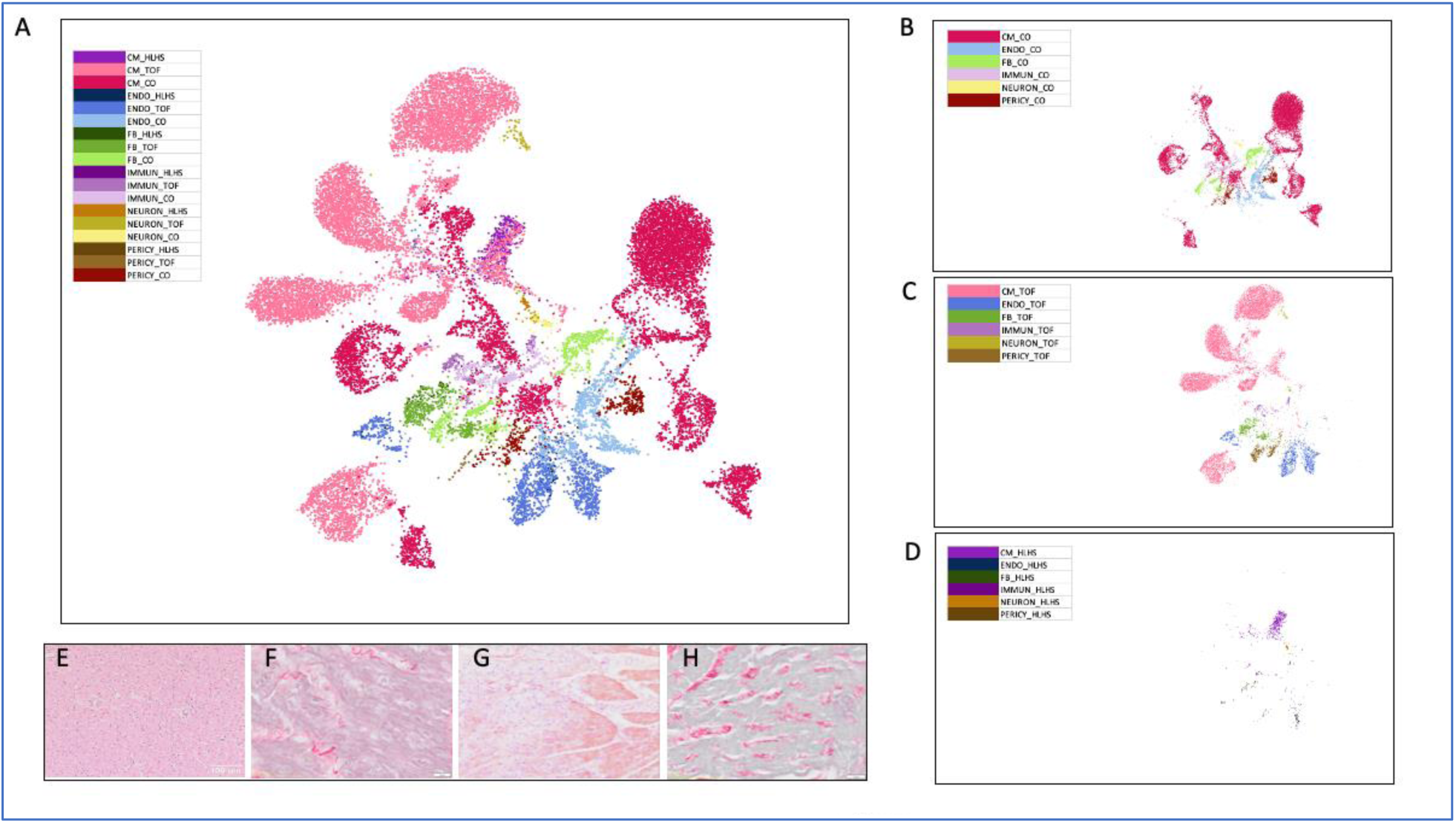
UMAP of all cells displayed with color coding in right ventricular myocardial samples in groups hypoplastic left heart syndrome (HLHS), tetralogy of Fallot (TOF) and Controls (CO), and histology pictures. A. UMAP of the 6 main cell types in the three groups (HLHS, TOF and CO) B. UMAP of the six main cell types in CO C. UMAP of the six main cell types in TOF D. UMAP of the main six cell types in HLHS E. Hematoxylin and eosin (HE) staining of right ventricular myocardial samples of TOF F. Picrosirius red (PSR) staining of right ventricle of TOF G. HE staining of HLHS H. PSR staining of HLHS CM: cardiomyocytes, ENDO: endothelial cells, FB: fibroblasts, IMMUN: immune cells, NEURON: neuronal cells; PERICY: pericytes; HLHS: Hypoplastic Left Heart Syndrome; TOF: Tetralogy of Fallot; CO: Controls.

### Gene expression profiles in different cell types compared to controls

For snRNA sequencing, HLHS1 and HLHS2 samples were pooled, due to low number of nuclei per samples. Significantly (p<0.01) fewer nuclei were found in HLHS samples than in TOF samples and controls (661 vs. 11,668 and 10,000). RNA-seq analysis identified 22 different subtypes of the six main cell types (cardiomyocytes [CMs], fibroblasts [FBs], pericytes, immune cells, neuronal cells, and endothelial cells [ECs]) in the right ventricular myocardial samples (Fig. 1).

Detailed differences in gene expression are listed in Supplementary Table S2. There were several similarities between the groups regarding pericytes, neuronal cells, and immune cells, but CMs, FBs, and ECs had obvious dissimilarities in gene expression between the groups (Fig. 2).

**Figure 2.**
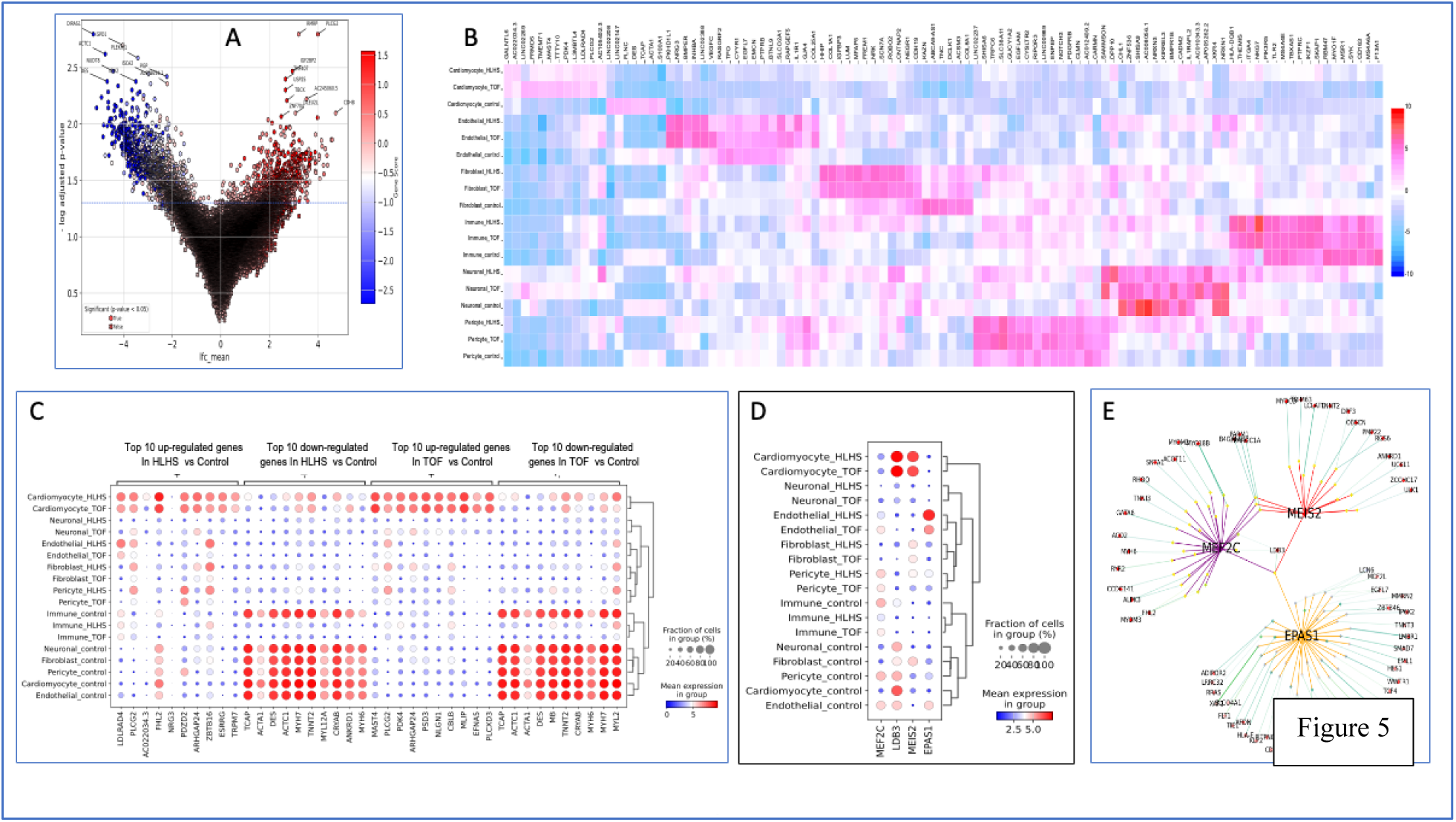
Highest de-regulated genes in the right ventricular samples of HLHS, TOF and controls. A. Volcano plot of all genes de-regulated in HLHS and TOF samples compared to controls. B. Heatmap of the top 100 de-regulated genes in the six main cell types in the three groups. C. Dot plot comparison of the differential gene expression between the groups. D. eRegulon showing the expression of the greatest significantly de-regulated genes across annotated cell types in both disease and control conditions. E. Visualization of the eRegulon for homeobox protein Meis2 (MEIS2), myocyte-specific enhancer factor 2C (MEF2C), and endothelial PAS domain-containing protein 1 (EPAS1) transcription factors, showing a dominant role of LIM domain binding 3 (LDB3).

A summarized comparison of the greatest de-regulated genes between the three groups (Fig. 2) revealed that, among the 4046 (347 genes in each HLHS nuclei, Suppl. Table S3) and 13,185 (12,369 genes in each TOF nuclei, Suppl. Table S4) genes significantly de-regulated in the HLHS and TOF samples compared to controls, respectively, the top over-expressed genes were wingless-type MMTV integration site family, member 9 (*WNT9*), EYA transcriptional coactivator and phosphatase 4 (*EYA4*), and troponin C1 (*TNNC1*). The greatest under-expressed genes were membrane protein CD36, thioredoxin-interacting protein (*TXNIP*), myoglobin (*MB*), and histone demethylase (*UTY*).

Clustering the genes in enhancer-driven gene regulatory networks (eRegulons), distinct signaling pathways were found, including actomyosin structure organization, early endosome membrane, cellular senescence, cardiac muscle contraction, dilated cardiomyopathy, and peroxisome proliferator-activated receptor (PPAR) (Fig. 2). Overexpression of myocyte-specific enhancer factor 2C (*MEF2C*), LIM domain binding (*LDB3*), and homeobox protein Meis2 (*MEIS2*) compared to controls was found in both HLHS and TOF samples. Endothelial PAS domain-containing protein 1 (EPAS1, known also as hypoxia-inducible factor-2alpha /HIF-2α/) was under-expressed in TOF. The CM-annotated clusters showed enrichment of MEF2C family motifs. Among the over-expressed genes, *MEIS2* was the most significant and is of particular interest due to its role in congenital defects. Visualization of the eRegulon for the MEIS2, MEF2C, and EPAS1 transcription factors showed a dominant role of LDB3, which is associated with cytoskeletal actin and muscle alpha-actin binding protein (Fig. 2).

In addition, *AC022034.3*, a glycolysis-related long non-coding RNA that is immune-active and associated with hypertrophic cardiomyopathy and infertility ^24^; *GALNTL6*, a protein-coding gene with a role in human fetal development and risk of CHD ^25^; and *L3MBTL4*, a protein-coding gene associated with multiple congenital defects ^26^ were significantly up-regulated in HLHS and TOF samples compared to controls (Fig. 2).

The top 50 up- and down-regulated genes are shown in Supplementary Figure S2 according to selected clusters of all cell types.

Violin plots of the greatest significantly de-regulated genes in cardiomyocytes are displayed in Supplementary Fig. S3.

For the HLHS samples, pathway enrichment analysis showed involvement in muscle contraction and extracellular matrix organization, with collagen formation and biosynthesis pathways, as well as several genes involved in anatomical structure development, cell adhesion, actin-myosin filament sliding, contractile muscle fibers, and metabolism pathways (Fig. 3, Supplementary Table S3).

**Figure 3.**
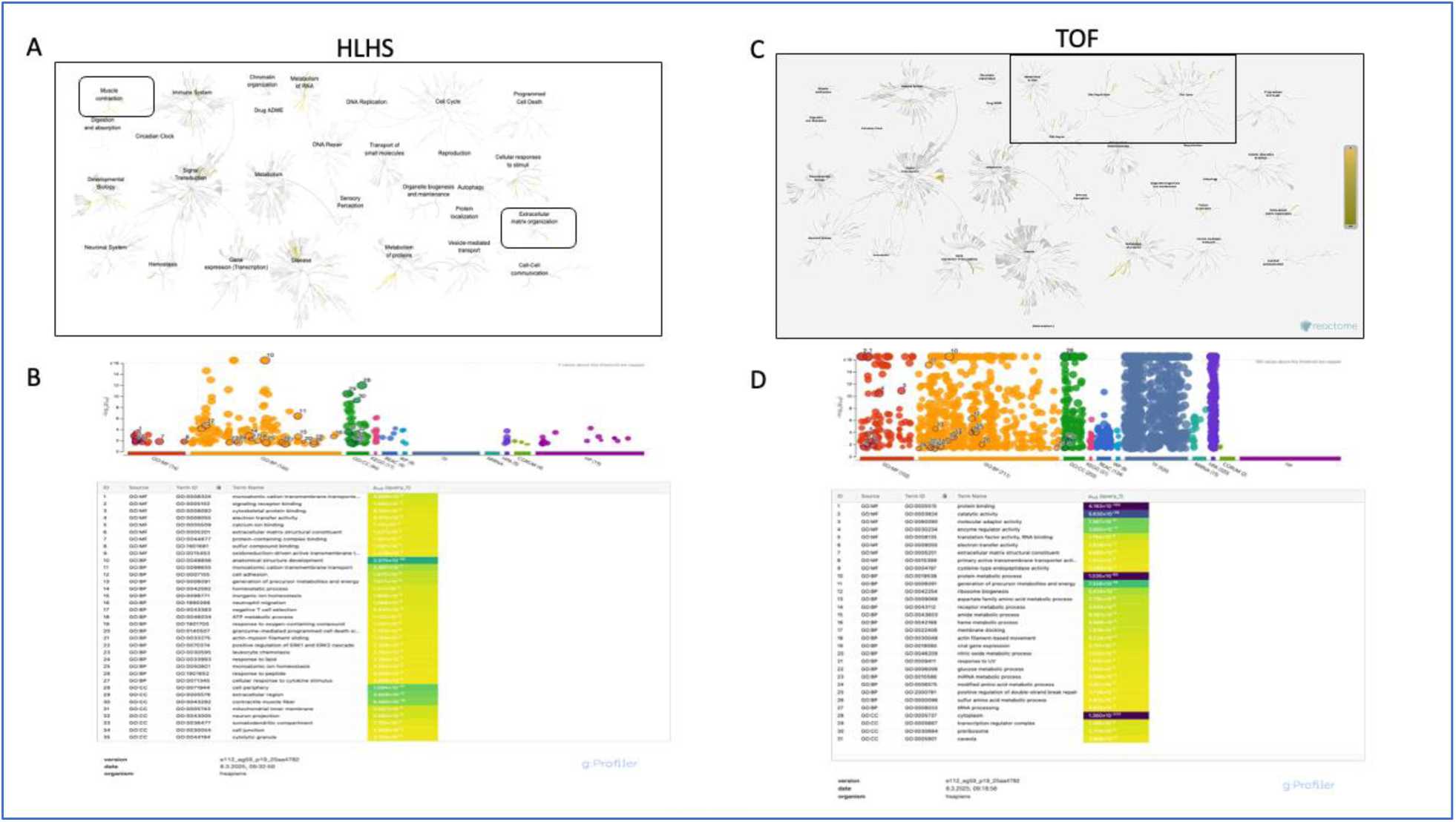
Reactome pathway and gProfiler analyses of the top 100 de-regulated genes in the hypoplastic left heart syndrome (HLHS) and tetralogy of Fallot (TOF) samples. A. Reactome pathway with integrated de-regulated genes in the pathways of HLHS B. Functional profiling of the 100 significantly de-regulated genes of HLHS C. Reactome pathway of significantly de-regulated genes of TOF samples D. Functional profiling of the 100 significantly de-regulated genes in TOF disease

In contrast, TOF samples showed involvement in cell cycle, DNA repair, DNA replication, and RNA metabolism pathway networks with several significantly de-regulated genes, including transcription factors and microRNAs (Fig. 3, Supplementary Table S4).

### Gene expression maps of the cell subtypes

Several cell subtypes were not present in significant amounts, especially in the HLHS samples (Fig. 4).

**Figure 4.**
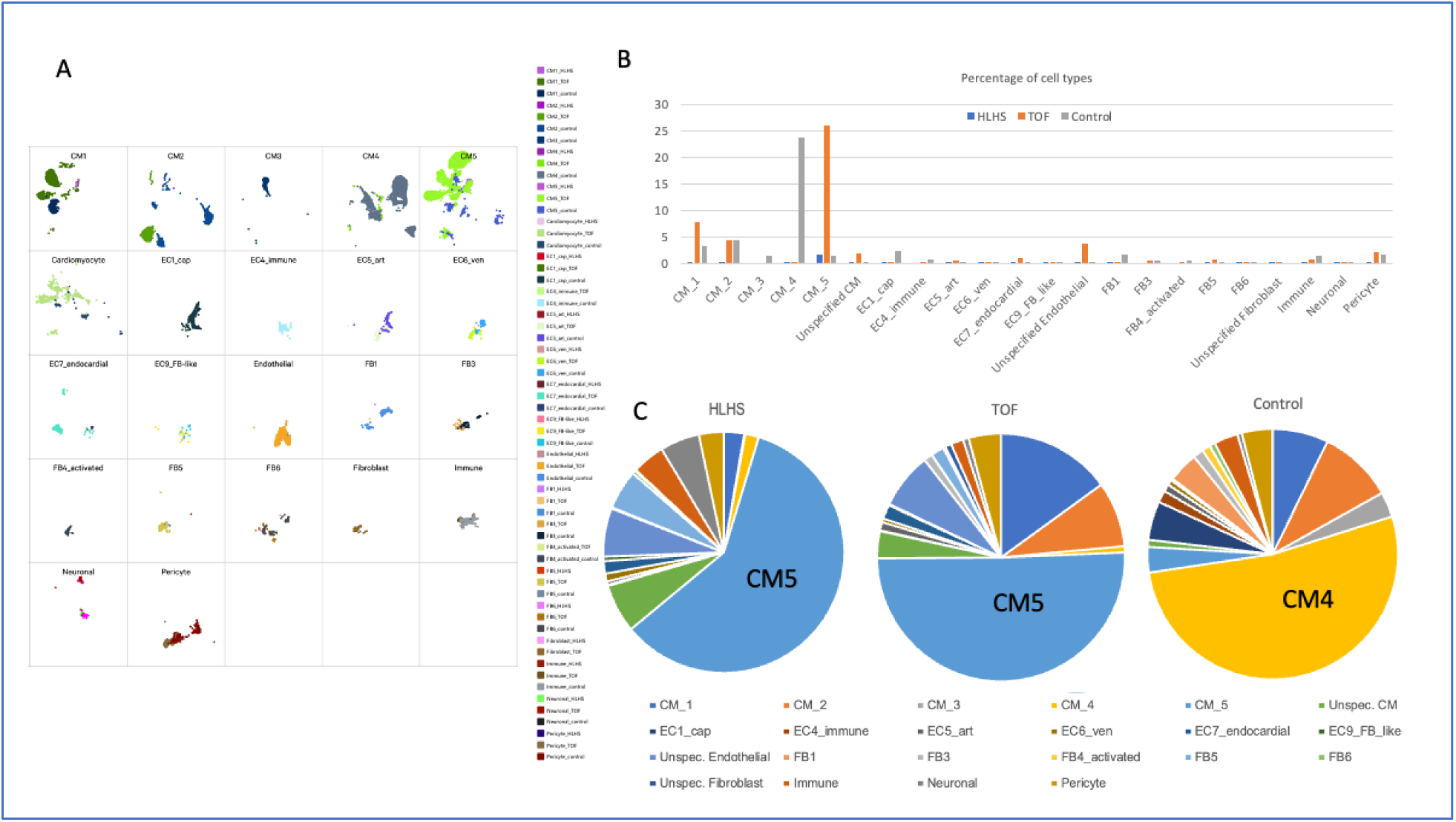
Distribution of the cell subtypes in the three groups. A. UMAP of all cell types in the three groups. B. Percentages of the cell subtypes. C. Bar graph of the cell subtype distributions. HLHS, hypoplastic left heart syndrome; TOF, tetralogy of Fallot; CO, controls; CM, cardiomyocytes; EC, endothelial cells; FB, fibroblasts.

The dominant cell type in HLHS and TOF samples was CM subtype 5 (CM5). Interestingly, the dominant CM subtype in the control group was CM4, and CM3 markers (FHL1+CNN1+MYH9; www.humancellatlas.org) were detectable only in the control samples. Other missing cell types in HLHS samples were subtypes of ECs and FBs: EC4 immune cells, FB3, and activated FB4.

The most plausible explanation for not detecting these cells is the small number of nuclei in the HLHS biopsy samples. However, CM3 could also not be annotated in the TOF samples. The right ventricular biopsy samples from HLHS, TOF, and controls contained 70.5%, 78.4%, and 76.4% CMs, respectively. In contrast, more FBs were found in HLHS samples than TOF and control samples (5.9% vs. 2.9% and 1.79%).

Considerable heterogeneity of gene expression was observed at the single-cell level. For example, pooling the TOF CM1 nuclei (n=1705), 7284 genes were significantly differentially expressed compared to controls, but only 419 (5.8%) significantly de-regulated genes were similarly expressed in each CM1 cell from the TOF samples. The mean similarity of the gene expression of pooled nuclei of the same cell subtype was 63.1% in the HLHS samples and 35.6% in the TOF samples. This genetic heterogeneity of the same cell phenotype may lead to further epigenetic modulation, leading to diverse clinical presentation.

### Gene profiling of the cardiomyocyte subtypes

Functional profiling of the significantly de-regulated genes in the five CM phenotypes (Fig. 5, Supplementary Table S5) revealed several clusters of genes differing between the three groups, including anatomical structure development, structural constituent of muscle, and cardiac muscle contraction.

**Figure 5.**
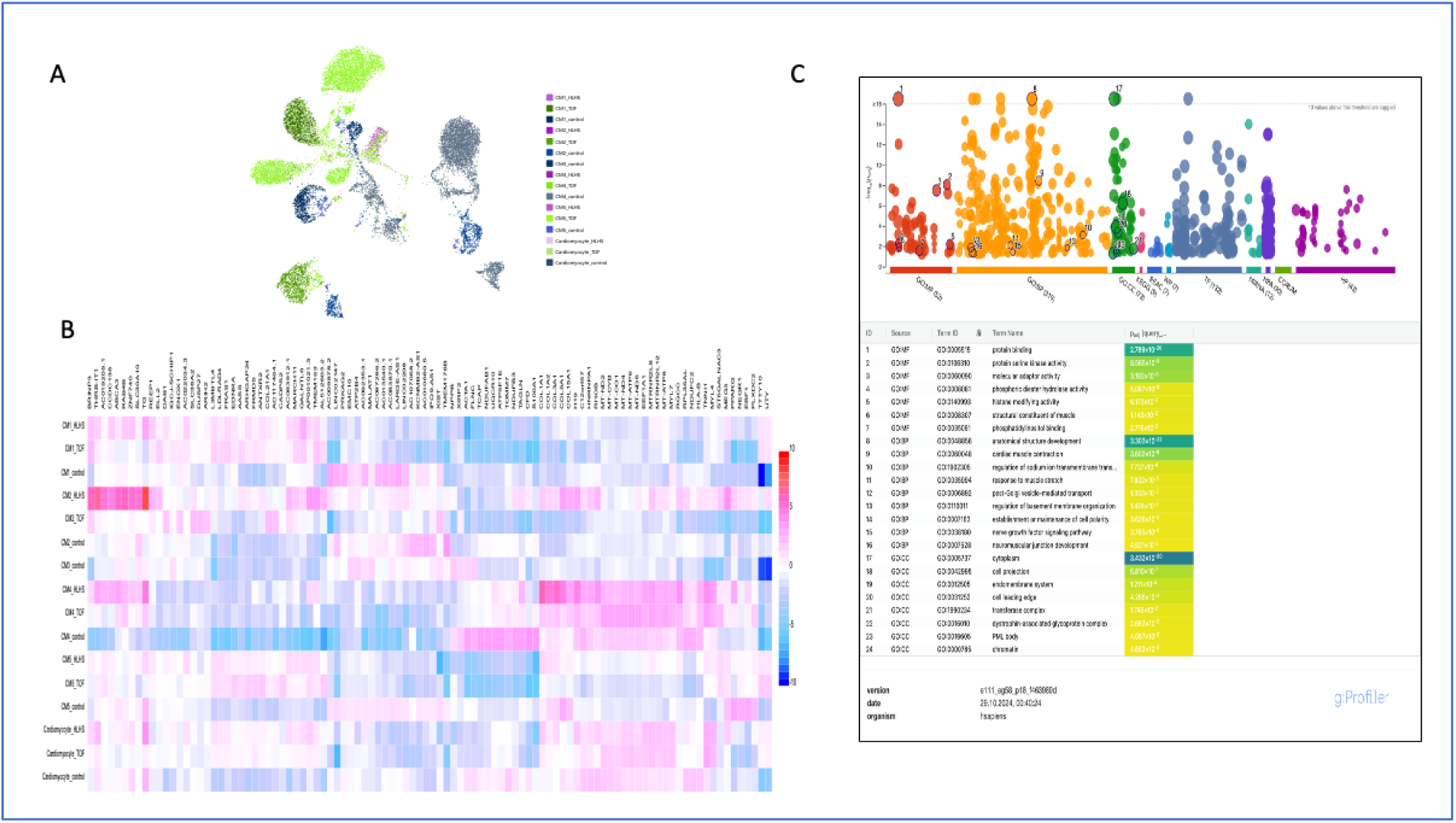
Analysis of the five cardiomyocyte subtypes in the hypoplastic left heart syndrome (HLHS), tetralogy of Fallot (TOF), and control myocardial samples. A. UMAP of the five types of cardiomyocytes in the three groups. B. Heatmap of the top 100 de-regulated genes in the different cardiomyocyte phenotypes (CM1-CM5). CM3 was found only in control myocardial samples. C. Functional profiling of the significantly de-regulated genes in the cardiomyocyte phenotypes.

Further downstream analyses of the CM subtypes are shown in Figures 6-8. Investigating CM1, HLHS samples contained few cells of this subtype, with 28 significantly de-regulated genes, 20 of which were de-regulated in each sample of CM1 from HLHS (Fig. 6, Supplementary Table S6). Some genes built the translational initial factor cluster, such as ubiquitin carboxyl-terminal hydrolase FAF-Y (*USP9Y*), which is responsible for protein integrity. No significantly down-regulated genes were found in CM1 from HLHS samples compared to controls.

**Figure 6.**
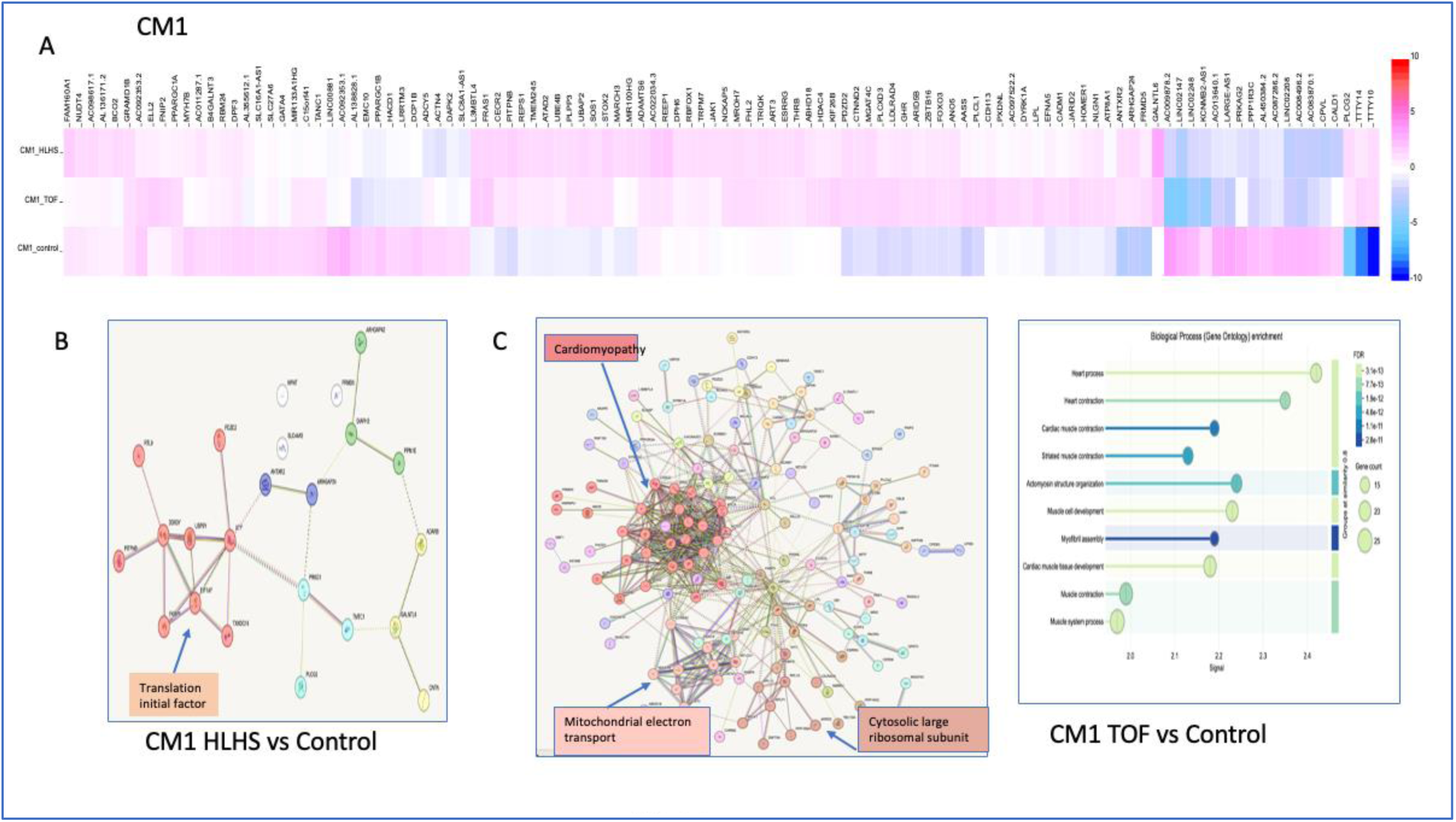
Significantly de-regulated genes in type 1 cardiomyocytes (CM1) in the three groups. A. Heatmap of the top 100 de-regulated genes in the three groups. B. Protein-protein interactions of significantly up-regulated genes in hypoplastic left heart syndrome (HLHS) vs. controls. C. Left, Protein-protein interactions displaying the 212 significantly de-regulated genes in CM1 between tetralogy of Fallot (TOF) and control samples. 23 genes with no connection to other genes were excluded. The three main clusters are displayed. Right, Functional visualization of Gene Ontology enrichment analysis of TOF vs. control. Significant differences were found regarding cardiac muscle development, muscle contraction, and actomyosin structure organization.

Interestingly, eight genes were expressed sporadically and not in all CM1 from HLHS samples, including nephronectin (*NPNT*), which is involved in organ development; protein kinase CGMP-dependent 1 (*PRKG1*), which is involved in aortic malformation; diaphanous related formin 3 (*DIAPH3*), which is associated with diverse congenital diseases; and muscular LMNA interacting protein (*MLIP*), which is related to myopathy and intellectual disorders.

The common significantly de-regulated genes in CM1 from TOF versus control samples (Supplementary Table S7) and the protein-protein interaction analysis with the three main clusters of genes (clustering by MCL cluster, inflation parameter 3) related to cardiomyopathy, mitochondrial electron transport, and cytosolic large ribosomal subunit are shown in Figure 6. Gene Ontology (GO) enrichment analysis found significant differences between TOF and controls in regard to CM1 expression of cardiac muscle development, cardiac muscle cell differentiation, cytoskeletal and actin-binding activities, and myofibril or sarcomere cellular components. Among the 2048 significantly up-regulated genes, 304 were overexpressed in each single nucleus. Clustering of all these overexpressed genes resulted in 25 having a role in muscle cell development. Most central genes were titin (*TTN*), myozenin2 (*MYOZ2*), myozemin1 (*MYOM1*), calsequestrin2 (*CASQ2*), which belong to intrinsic cardiomyopathy or myofibrillar myopathy.

The most relevant down-regulated genes of the CM1 phenotype in TOF were tenascin (*TNN*), myosin heavy chain 7 (*MYH7*), GATA binding protein 4 (*GATA4*), actin alpha 1 (*ACTA1*), and myosin light chain 2 (*MYL2*) of the myofibril assembly and cardiac muscle development clusters.

Similar to CM1, a small number of CM2 nuclei were detected in the HLHS samples. The CM2 gene expression of HLHS samples was not altered compared to controls. In contrast, CM2 gene expression analysis revealed significantly altered expression of several genes in TOF samples versus controls (Fig. 7, Supplementary Table S8). The genes were clustered in oxidative phosphorylation, cytoplasmic translation, myofibril assembly, and muscle proteins. The biological process GO enrichment analysis is presented in Figure 7.

**Figure 7.**
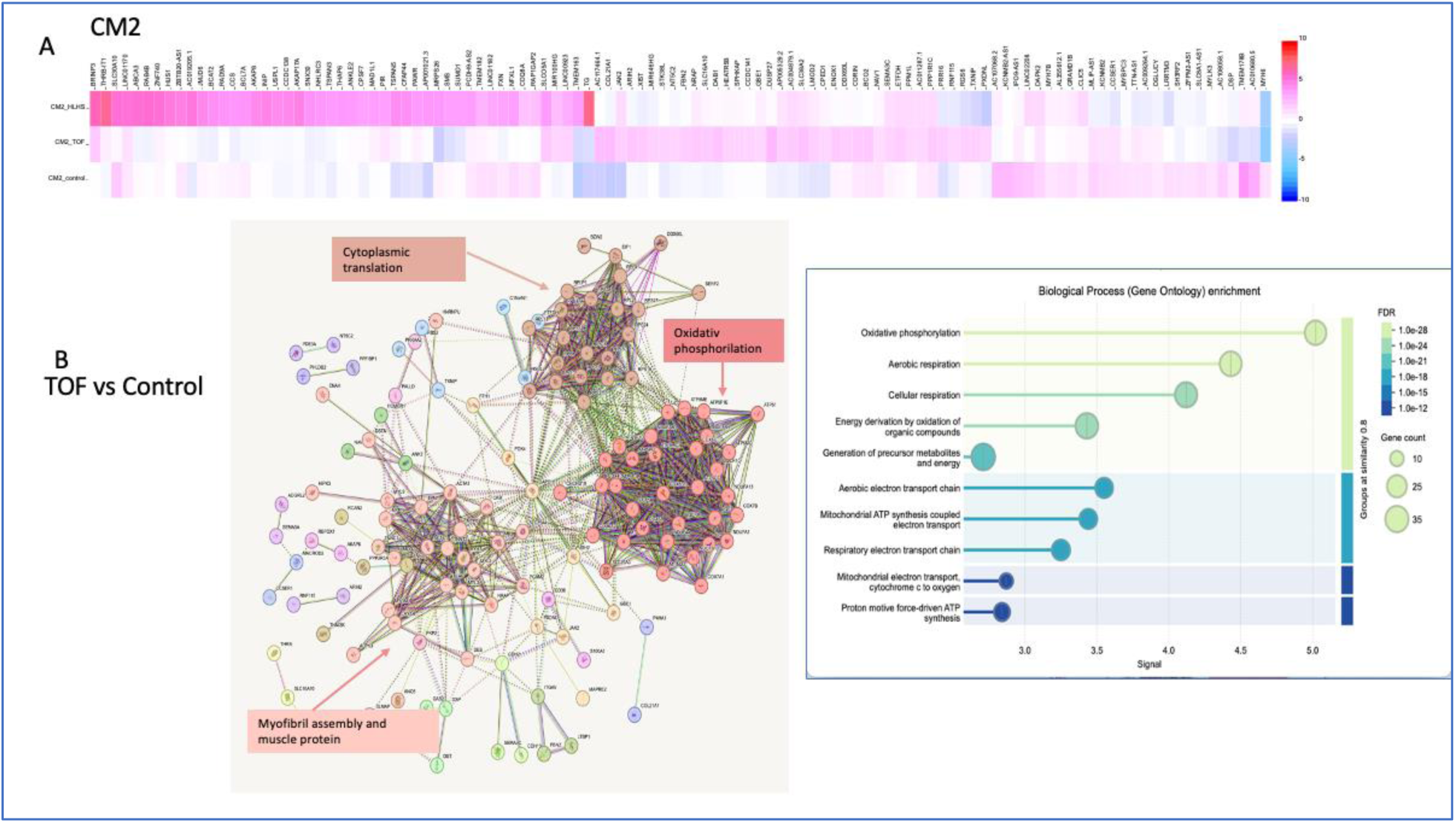
Analysis of the differentially de-regulated genes in type 2 cardiomyocytes (CM2) in the three groups. Hypoplastic left heart syndrome (HLHS) samples only contained one nucleus. A. Heatmap of the top 100 significantly de-regulated genes. B. Protein-protein interactions displaying the significantly de-regulated genes between tetralogy of Fallot (TOF) and controls after excluding genes with no connection to other genes. The three main clusters are displayed. C. Functional visualization of the Gene Ontology enrichment analysis of TOF vs. controls. Significant differences were found regarding cardiac metabolism.

The most important cluster of the significantly up-regulated CM2 genes in TOF samples was RAR-related orphan receptor A (RORA), which plays a central regulatory role in cellular differentiation, embryonic development, and organogenesis. The down-regulated CM2 genes in the TOF samples belonged to cardiac tissue morphogenesis, including *MYH7*, *MYL* family, *TNN* family, *LDB3*, *GATA4*, *ACTC1*, and tropomyosin (*TPMA*).

Interestingly, neither the HLHS nor the TOF samples contained CM3. Supplementary Figure S3 displays the greatest up- or down-regulated CM3 genes with the GO enrichment analysis indicating a role in cardiac muscle tissue and cell development, actin filament-based processes, and actomyosin structure organization (Supplementary Table S9).

The CM4 gene expression profiles were similar in all three groups (Supplementary Fig. S4, Supplementary Tables S10 and S11).

In the HLHS samples, CM5 expressed >100 significantly up- and down-regulated genes (Supplementary Table S12). The largest cluster of up-regulated genes was tyrosine kinase inhibitor, postsynaptic density and cell adhesion, which are associated with several genetic phenotypes of congenital diseases (e.g., autism, neurogenic disorders, or bone dysmorphogenesis). The greatest number of down-regulated genes were clustered into extracellular matrix organization, oxidative phosphorylation, and cardiac tissue morphogenesis, including *COX5B*, *COX7C*, ATP gene family (e.g., *ATP5PO, ATP5MD, ATP5PF*), NADH dehydrogenase family, *MYH7, MYL2, MYL3, MYL7, TNN2, TNN3, ACTC1*, and creatine kinase M (*CKM*) (Fig. 8).

**Figure 8.**
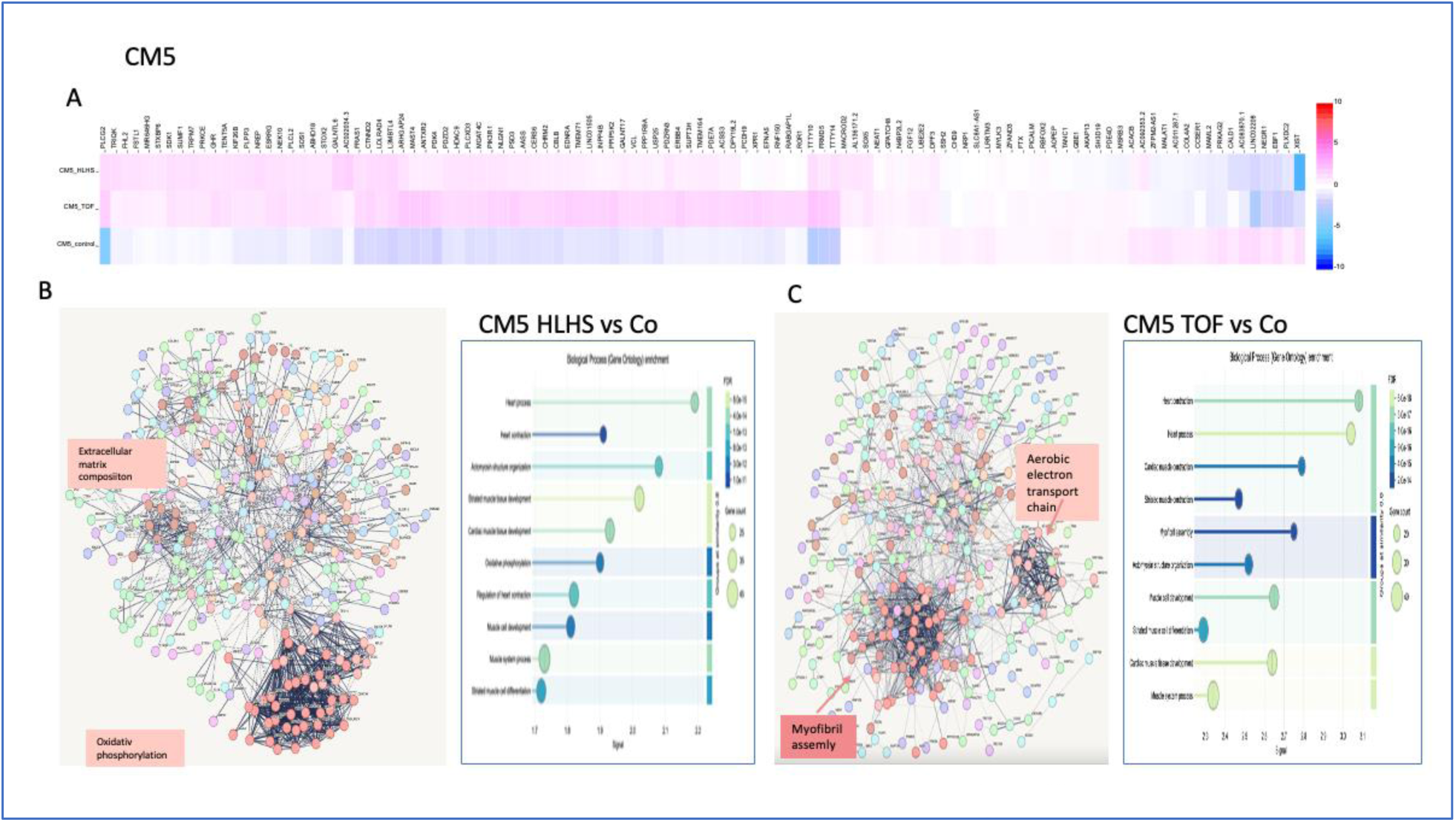
Analysis of the differentially de-regulated genes in type 5 cardiomyocytes (CM5) in the three groups. A. Heatmap of the top 100 significantly de-regulated genes. B. Left, Protein-protein interactions displaying the significantly de-regulated genes between hypoplastic left heart syndrome (HLHS) and controls. The two most important clusters are shown. Right, Functional visualization of Gene Ontology enrichment analysis, showing a role in heart process and actomyosin structure organization. C. Protein-protein interactions displaying the significantly de-regulated genes between tetralogy of Fallot (TOF) and controls. The two main clusters are displayed. Right, Functional visualization of Gene Ontology enrichment analysis revealed significant differences between TOF and controls regarding cardiac muscle contraction, muscle cell development, myofibril assembly, and actomyosin structure organization.

Similarly, 384 genes were de-regulated in the TOF samples compared to controls (Supplementary Table S13). Figure 14 shows the two main clusters of these genes belonging to myofibril assembly and aerobic electron transport chain. Biological process GO enrichment analysis revealed genes in the heart muscle contraction and muscle cell development pathways, among others.

### Profiling differential gene expression in the other cell subtypes

The proportion of nuclei from non-CM subtypes in all groups was <10%; therefore, we performed combined biological process GO enrichment analyses of the significant gene expression profiles (Supplementary Figures S5-S8).

#### Endothelial cell subtypes

Gene expression was similar in the three groups of the EC subtypes such as EC1 capillaries, EC4 immune, and EC9 FB-like.

EC5 arterial gene expression was similar in HLHS and controls. However, TOF samples had 96 significantly up-regulated EC5 genes, compared to EC5 genes of controls, including vimentin (*VIM*), which is found in mesenchymal cells, and endothelial PAS domain-containing protein 1 (*EPAS1*), a regulator of vascular endothelial growth factor (VEGF) expression, blood vessel development, and endothelium formation that belongs to the angiogenesis (Supplementary Fig. S5, Supplementary Table S14). We identified 126 significantly down-regulated genes, building three main clusters: oxidative phosphorylation (COX family, NDUF family, and ATP family), cardiac tissue morphogenesis (triadin [*TRDN*], Ankyrin repeat domain-containing protein 1 [*ANKRD1*], with a role in EC activation), and cytoplasmic translation with several ribosomal genes (RPS family).

Only five genes with the role of cellular defense against viral entry (interferon-induced transmembrane protein 3 [*IFITM3*]) and antigen processing (HLA class II histocompatibility antigen gamma chain [*CD74*]) were up-regulated in the EC6 venous subtype from HLHS samples (Supplementary Table S15), but several up- and down-regulated genes were found in TOF samples compared to controls (Supplementary Table S16). For example, the Hippo pathway with components of epidermal growth factor (EGFR) and VEGF-1, and the HIF1-alpha and VEGF ligand receptor pathways (*EPAS1*, insulin receptor subunit alpha [*INSR*]) were up-regulated, playing a role in embryonic vasculature development and the regulation of angiogenesis (Supplementary Fig. S5). Most relevant down-regulated genes were classified into oxidative phosphorylation (COX family, NDUF family, and ATP family motifs) and aerobic electron transport chain (COX family, NDUF family, and cytochrome [CYC]) clusters.

Differential gene expression was observed in the EC7 endocardial cell subtype for both HLHS and TOF samples compared to controls (Supplementary Tables S17, S18). The most relevant up-regulated genes encode the extracellular matrix components versican core protein (*VCAN*) and collagen alpha-2(I) chain (*COL1A2*). The two main clusters of down-regulated genes were myofibril assembly and oxidative phosphorylation, including *MYH*, *MYL* family, *TNN* family, *MYOZ22*, and *MB* (Supplementary Fig. S5).

#### Fibroblast subtypes

FB3 and FB4 in TOF samples and FB5 in both HLHS and TOF samples differed significantly from controls (Supplementary Fig. S6, Supplementary Tables S19-S22).

Forty-three genes were up-regulated in FB3 from TOF samples and belonged to the extracellular matrix proteoglycan and collagen fibril organization clusters (COL family: *COL3A1*, *COL1A1*, *COL1A2*; *VIM*). In addition, several cardiac muscle morphogenesis (*TNN*, *MYH* and *MYL* families, desmin [*DES*], alpha-crystallin B chain protein [*CRYAB*], *ACTC1*, *TCAP*) and oxidative phosphorylation (*COX*, ATP and NDUF gene families) genes were significantly down-regulated compared to controls Supplementary Fig. S6).

Thirty-six genes were up-regulated in FB6 from TOF samples and belonged to the collagen biosynthesis and extracellular matrix organization clusters (COL family), as well as insulin-like growth factor (*IGFBP3* and *IGFBP5*). Twenty genes were down-regulated, including those with a role in striated muscle contraction (ACT family, *TNNT2*, *ACTA1*, *CRYAB*) (Supplementary Fig. S6).

More than 100 genes were up-regulated in FB5 from HLHS samples compared to controls and were classified into the extracellular matrix and collagen fibril organization and collagen binding clusters (COL family, such as *COL3A1, COL5A2, COL6A1*; fibrillin-1 [FBN-1]; fibulin-5 [*FBLN5*]), with a role in the structural organization of microfibrils of the extracellular matrix, elastic fibers, and connective tissues. Further up-regulated genes were A disintegrin and metalloproteinase with thrombospondin motifs 5 (*ADAMTS5*) and matrix metallopeptidase 2 (*MMP2*), which plays an important role in connective tissue organization and remodeling of the vasculature, angiogenesis, tissue repair, tumor invasion, and inflammation. Up-regulated genes tenascin-X (*TNXB*) and decorin (*DCN*) are involved in cell-extracellular matrix interactions and acceleration of collagen fibril formation (Supplementary Fig. S6). We did not find significantly down-regulated genes in FB5 from HLHS samples.

In TOF samples, the up-regulated FB5 genes were involved in extracellular matrix and collagen fibril organization and collagen binding, including the COL family (e.g., *COL3A1, COL5A2, COL6A1*), *FBN1*, *FBLN5*, protein-lysine 6-oxidase (*LOX*), *FN1*, microfibrillar-associated protein 2 (*MFAP2*), and serpin H1 (*SERPINH1*) (Supplementary Fig. S6).

#### Immune cell subtype

The significantly de-regulated genes of immune, and neuronal cells are displayed in Supplementary Fig. S7, and listed in Supplementary Tables S23-S26.

Immune cells from the HLHS samples had several up-regulated genes compared to controls that clustered into antigen processing and the presentation of peptide antigen and the MHC protein complex with HLA class I histocompatibility antigens A, B, C, and alpha chains (*HLA-A, HLA-B, HLA-C*); the antigen-specific T cell immune response; antigen-presenting major histocompatibility complex class I (*MHCI*) or CD74 (cell surface receptor for the cytokine macrophage migration inhibitory factor [MIF]); and the Baraitser-Winter syndrome pathway, which is involved in the clinical phenotype of multiorgan genetic defects in mainly the brain and face. CRYAB, a member of the heat shock proteins that is associated with cardiac abnormalities, was down-regulated (Supplementary Fig. S7).

Gene expression by immune cells from the TOF samples was partially similar to that of the immune cells from the HLHS samples, but the main cluster was the phosphatidylinositol signaling system (PTEN-tenascin, 1-phosphatidylinositol 4,5-bisphosphate phosphodiesterase gamma-2 [*PLCG2*] transmembrane signaling with the Ikaros gene [*IKZF1*]) that plays a role in the development of lymphocytes, B cells, and T cells, and in lymphocyte and T-cell activation. The down-regulated genes belonged to the oxidative phosphorylation cluster: COX family, NDUF family, and ATP family (Supplementary Fig. S7).

#### Neuronal cell subtypes

The neuronal cell subtypes in HLHS samples had several up-regulated genes compared to controls, including contactin associated protein 1 and 2 (*CNTNAP1* and *CNTNAP2*), the nerve impulse protein involved in postsynaptic cell membrane molecular organization and protein-protein interactions at synapses, as well as the longitudinal and radial organization of myelinated axons and nerve development. No significantly down-regulated genes were observed.

Up-regulated genes in neuronal cells from the TOF samples (Homer protein homolog 1 [*HOMER1*] and neuroligin-1 [*NLGN1*]) belonged to neurotransmitter-gated ion channel clustering, post-synapsis organization, neuronal cell signaling, and synaptic signal transmission. Down-regulated genes (COX family, NDUF family, RPS family, and ATP family) in the TOF samples were involved in oxidative phosphorylation, the aerobic electron transport chain, and mitochondrial metabolism (Supplementary Fig. S7).

#### Pericyte subtypes

Compared to controls, the pericyte subtypes from HLHS samples overexpressed basement membrane and vascular inflammation genes (*SPARC*, thrombospondin-4 [*THBS4*]), which are involved in adaptive pressure overload regulation to avoid myocardial stress, and fibronectin (*FN1*) and Fraser extracellular matrix complex subunit 1 (*FRAS*), which play a role in cell adhesion and migration processes, including embryogenesis and organogenesis (Supplementary Fig. S8). There were no significantly down-regulated genes in the HLHS samples (Supplementary Table S27).

In the TOF samples, several genes were up-regulated in pericytes compared to controls. These genes are members of the signal transduction network, including epidermal growth factor receptor (*EGFR*), *RORA*, and *FOXO3*. Down-regulated genes (RPS family: *RPS9, RPS16, RPS3A*; RPL family: *RPL9, RPL12, RPL24*; and *MYH* and *MYL* families) pertained to cytoplasmic translation, ribosome biogenesis, and myofibril assembly (Supplementary Fig. S8, Supplementary Table S28).

## Discussion

Here, we provided novel insights into altered gene expression patterns and pathways in myocardial samples from patients with HLHS and TOF, which are relevant to pathological cardiogenesis in CHDs. We performed primarily exploratory analyses using several unsupervised techniques, including differential gene expression, gene set enrichment, clustering, and dimensionality reduction. We also integrated third-party control samples, annotated the cell types, and constructed an eRegulon network. This exploratory phase has helped identify patterns and differences that are valuable for generating new hypotheses.

We identified >12,000 transcriptomes that are significantly de-regulated in right myocardial nuclei from 22 different cell clusters in HLHS and TOF compared to controls. Further bioinformatic tools revealed MEIS2, MEF2C, and EPAS1 transcription factors, with a dominant role of LDB3, which interacts with myozenin family motifs and is associated with cardiomyopathy, myofibrillar myopathy, heart development, and cardiac chamber structural anomalies.^27,28,29^ Functional clustering of the significantly de-regulated genes explored cell-nuclei cluster-specific dysregulation of heart development processes, metabolic abnormalities, and several other pathological pathways in cardiogenesis and cardiac structure organization. We also observed great heterogeneity in the gene expression of the cellular subtypes, which may explain the diverse clinical phenotypes within disease entities among CHDs.^30,31,32^

Relatively few nuclei were found in HLHS samples, in line with the histological findings in the same myocardial tissue. However, the multiome process allows exploratory analysis of a single nucleus, uncovering sufficient information at the single-cell resolution.^33^

Due to the difficulty obtaining pediatric cardiac tissue for research purposes, only a couple of comparative analyses exist in the literature. Hill et al. performed single-nucleus RNA-seq of nine pediatric samples and four controls.^34^ There are several important differences between their methodologies and ours. First, we clustered the CMs into five phenotypes, whereas Hill et al. used three. Second, we identified 22 subtypes of the six main cell types compared to the 14 subtypes identified by Hill et al. Third, we did not pool the nuclei of HLHS and TOF samples, and we had homogenous CHD entities from patients of similar age. We did not mix the nuclei of other CHDs or from different ages of patients for the analysis. However, we found major similarities with Hill et al.’s results regarding the de-regulated genes and signaling networks, such as FOXO and CRIM1 in CMs.^34^ Furthermore, we found significant gene alterations in the non-cardiomyocyte cell types too, highlighting the complex genetic processes in congenital heart diseases.

Nicin et al. described single-cell RNA-seq results in children with dilated cardiomyopathy and characterized the CMs into three subtypes, similar to Hill et al. ^35^ Single-cell RNA-seq data have also been reported for myocardial samples during the developmental stage, mostly in small animals, or CM-derived induced pluripotent stem cells (IPS) or CMs in cell culture. ^4, 30,36,37,38^ Other studies have performed histological analysis of human heart samples from patients ^39^ or fetuses with CHDs ^40^, or whole genome sequencing in family members of patients with CHDs. ^41^ Asp et al. performed spatiotemporal gene expression analysis in different developmental stages of CHDs and identified Myoz2 and Fabp3-enriched CMs, with time-dependent temporal specificity of several other cell types.^42^ Aguayo-Gómez identified copy number variations in 52 TOF patients and revealed *TBX1* deletion in one patient ^43^, whereas whole exome sequencing paired with microarray analysis suggested a central role of a de novo variant in *ROBO1*. ^44^ Genetic features of CHDs were also summarized in several reviews ^4,45,46,47,48,49,50^, highlighting the diverse proposed mechanisms of the genotypic and phenotypic abnormalities resulting in CHDs. Similar to previous studies, in the present study, different cardiac cell types had overlapping gene signatures, and the common feature of CM and non-CM nuclei were the cell nuclei-specific genetic fingerprints with de-regulation of genes involved in cardiac structure development, myofibril structure, and oxidative phosphorylation. Our downstream analysis revealed dysregulated genes in translation initiation in CM1 from HLHS; cardiomyopathy and mitochondrial electron transport pathway genes in CM1 from TOF; myofibril assembly, oxidative phosphorylation, and aerobic metabolism in CM2 and CM5 from TOF; and extracellular matrix organization transcripts in CM5 from HLHS. Our study surpasses the previous studies with a clear view of the de-regulated genes and pathways, allowing drug targeting and further research on the treatment of CHDs.

Our study has several limitations. In contrast with previous publications, we did not find clearly annotated smooth muscle cells (SMCs). CALD1 seems to be the only marker of SMCs that is significantly expressed; it is expressed evenly across all cells, overlapping other markers, which does not allow for clear annotation of SMCs. There were only few vessels in the samples (very low EC of arterial origin in HLHS samples and a small bit more in TOF samples), and the SMC gene expression overlaps with EC-art.

Annotation of cell subtypes becomes increasingly challenging at higher resolution, as finer distinctions are often obscured by a low signal to noise ratio caused by different technical and biological artifacts, such as multiplets and dropout phenomenon, which are limitations of sequencing technologies and single-nucleus RNA-seq. In addition, supervised cell annotation models are unable to identify and annotate uncharacterized cell types or states not present in the reference. Furthermore, disease-related changes in gene expression can entirely alter the transcriptomic profile of a cell.

Biopsy probes are snapshots of gene expression, missing the dynamic changes in nuclear and cellular gene expression. We analyzed the right ventricular tissue, though the cardiac anatomical structure also includes the atria, vessels (from large or medium-sized to capillaries), interatrial or intraventricular septa, electrical conduction systems, and heart valves. As both HLHS and TOF are associated with defects at several cardiac levels, a complex analysis including these structures is desirable.

The actual gene expression of a cell is influenced by pathological environmental factors, medication, stress, and sampling methods ^51^, but these limitations can be overcome by pooling multiple samples from several individuals. However, each pediatric myocardial biopsy or tissue is subjected to routine histology for diagnosis or disease staging and for individual therapy purposes; only a minority of the rest of the samples might be available for research.

In conclusion, we characterized 22 cell subtypes in six main cell groups in myocardial biopsies obtained from pediatric patients with HLHS or TOF and compared them to controls using single-nucleus RNA-seq. We revealed substantial heterogeneity of the same cell cluster, explaining the diversity of clinical phenotypes in CHDs. We found that CM3 was only present in control samples, and that the main CM phenotype was CM5 in HLHS and TOF and CM4 in controls.

We explored >12,000 differentially regulated genes in HLHS and TOF, with main pathways belonging to cardiac development, heart structure, oxidative phosphorylation, metabolic abnormalities, and dysregulated extracellular matrix pathways. Further exploration of the genetic basis of CHDs is warranted, with clear signals of druggable targets.

## Supplementary file

### Supplementary Figures

**Supplementary Figure S1.**
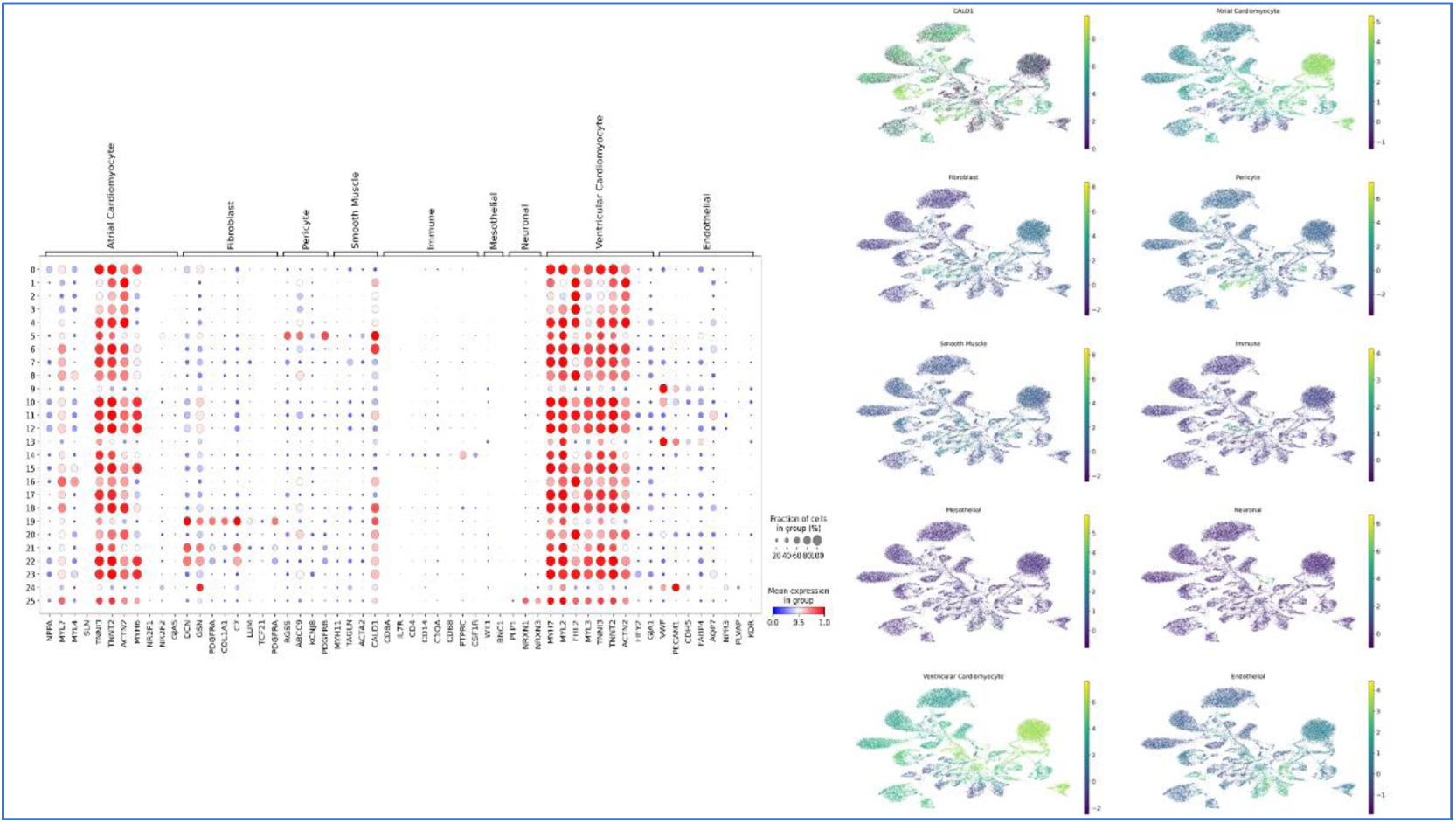
No smooth muscle cells (SMCs) were annotated because there was no particular cell cluster/group with higher expression in SMC marker genes. CALD1 seems to be the only marker of SMCs with significant expression. It was expressed evenly across all cells, overlapping other markers.

**Supplementary Figure S2.**
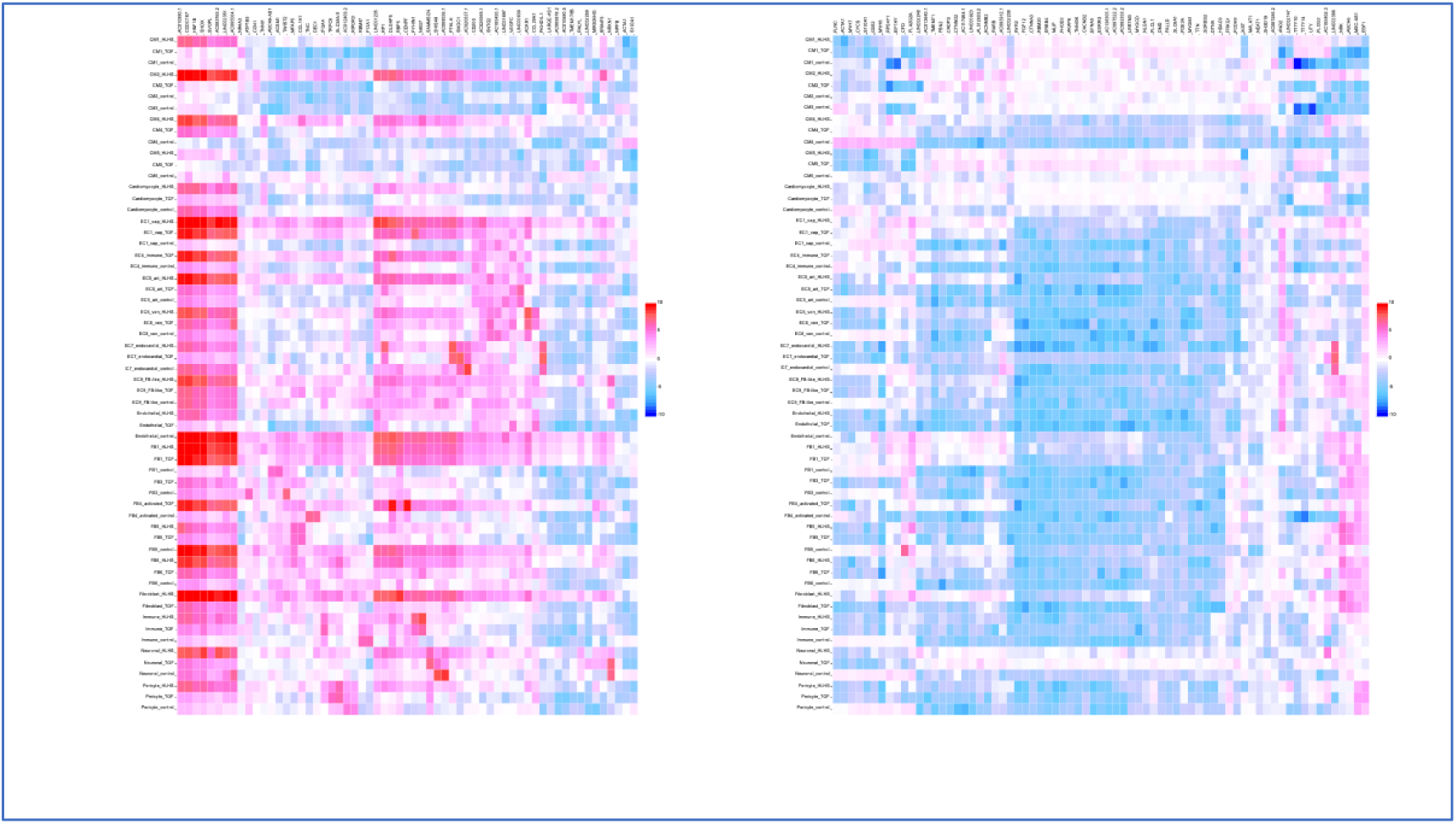
Top 50 up-regulated (left) and down-regulated (right) genes according to selected clusters of all cell types. Selection was made based on the highest p-value. The p-values were adjusted by Benjamini-Hochberg correction of multiple testing.

**Supplementary Figure S3.**
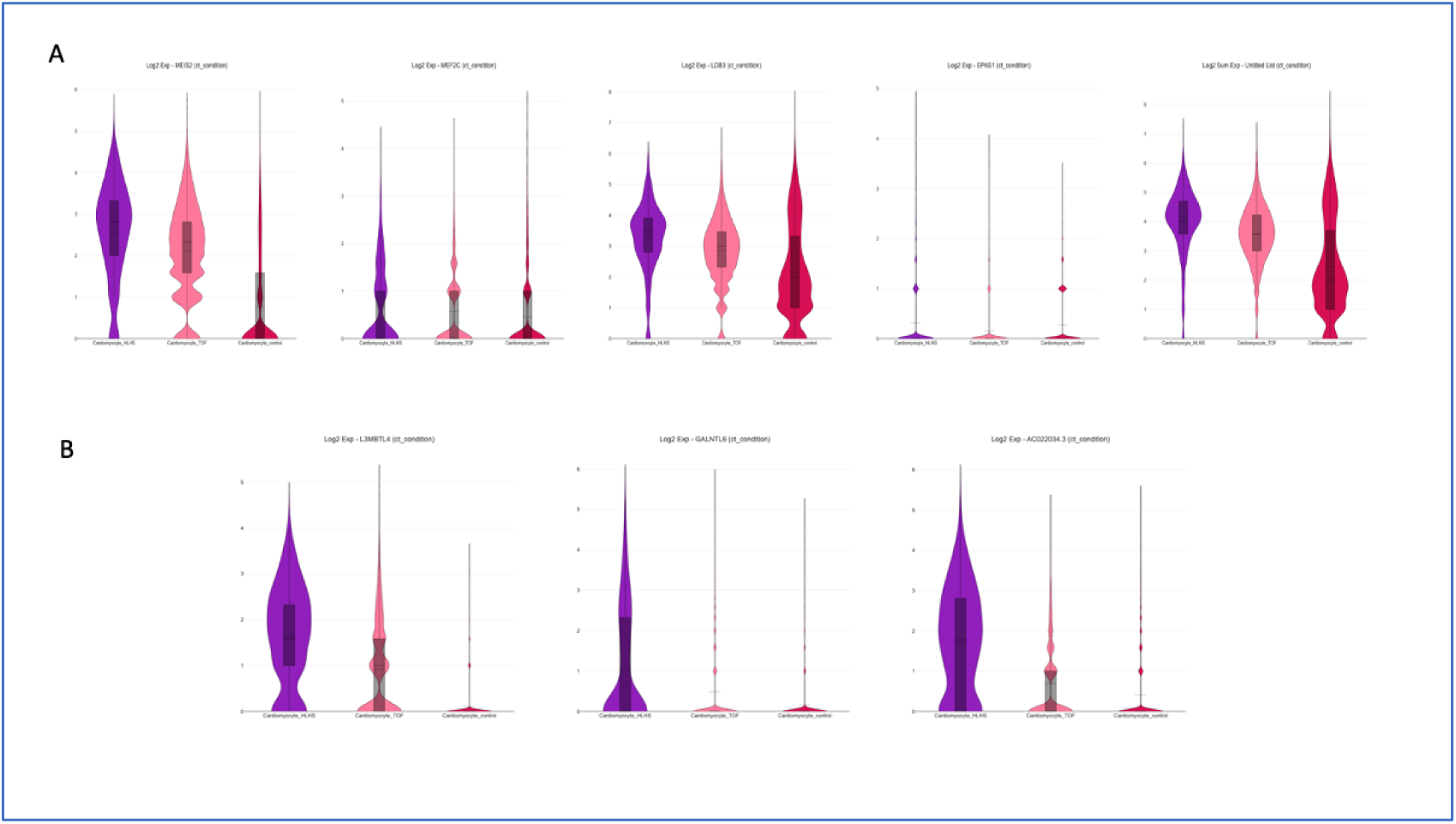
Violin plots of the greatest significantly de-regulated genes in cardiomyocytes. A. Violin plots of the singular and combined expression of the four greatest de-regulated genes determined by the eRegulon analysis including all cell types in the biopsy samples. B. Violin plots of the three greatest significantly de-regulated genes in cardiomyocytes.

**Supplementary Figure S4.**
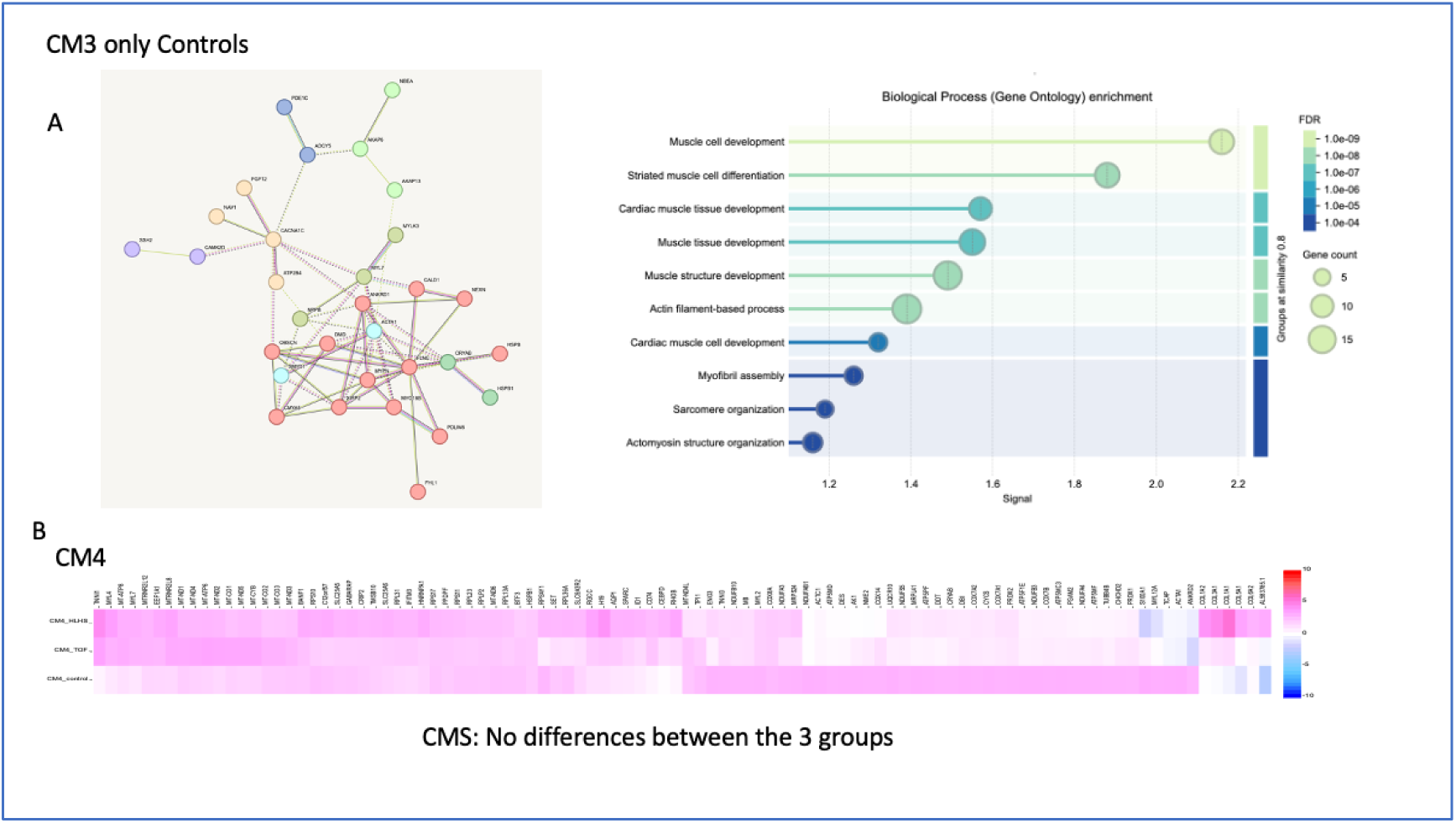
Top 50 and 100 de-regulated genes in cardiomyocyte subtypes 3 (CM3) and 4 (CM4). A. CM3 was found only in control myocardial samples. The top 50 up-or down-regulated genes in connection with the protein-protein interaction network and the functional enrichment analysis are shown. B. No differences were found between the three groups regarding gene expression in CM4.

**Supplementary Figure S5.**
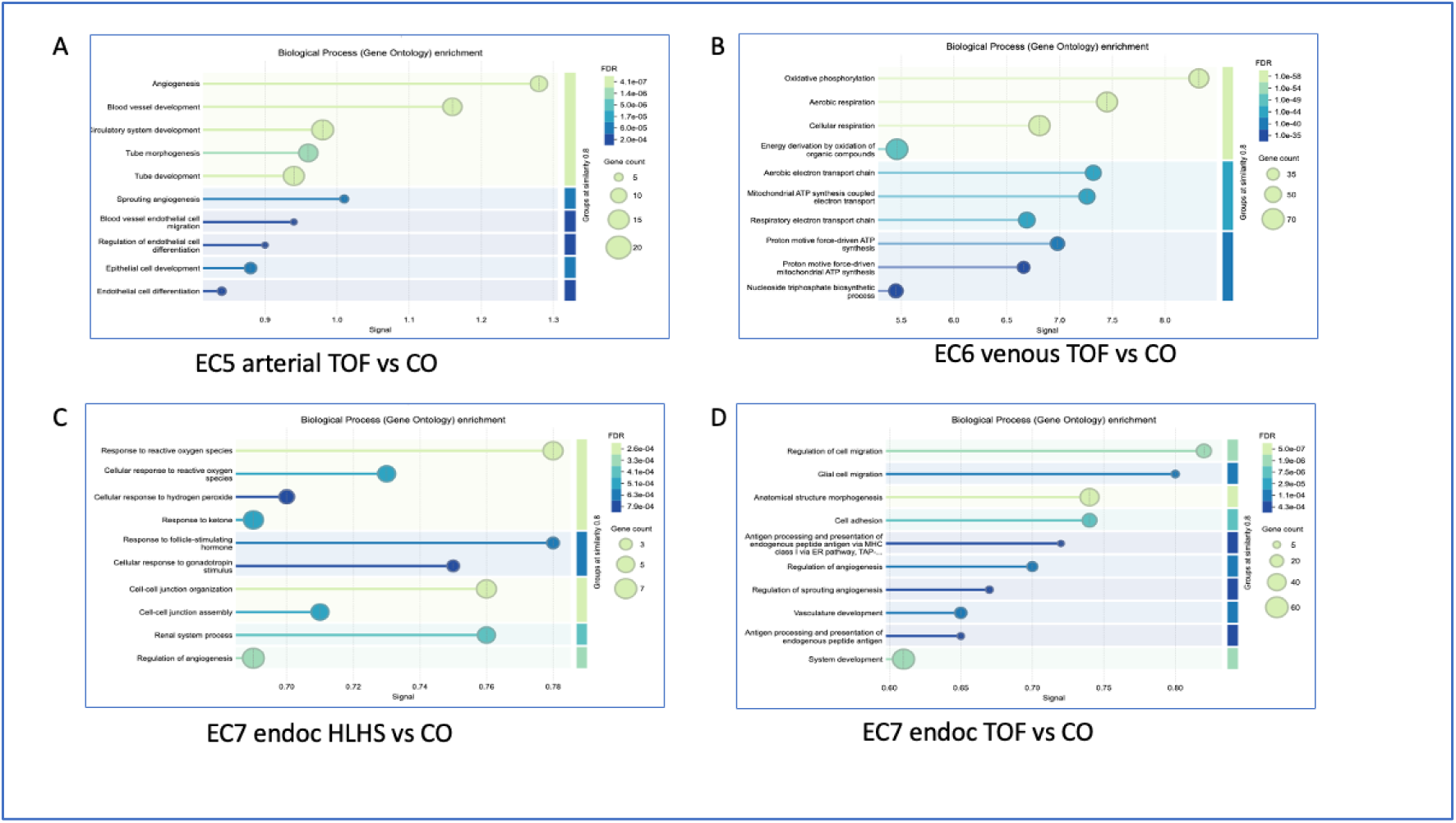
Gene Ontology enrichment analysis of the significantly de-regulated genes in endothelial cell (EC) subtypes from the pediatric cardiac biopsy samples. A. EC5 arterial in tetralogy of Fallot (TOF) vs. controls (CO). B. EC6 venous in TOF vs. CO. C. EC7 endocardial (endoc) in hypoplastic left heart syndrome (HLHS) vs. CO. D. EC7 endocardial in TOF vs. CO.

**Supplementary Figure S6.**
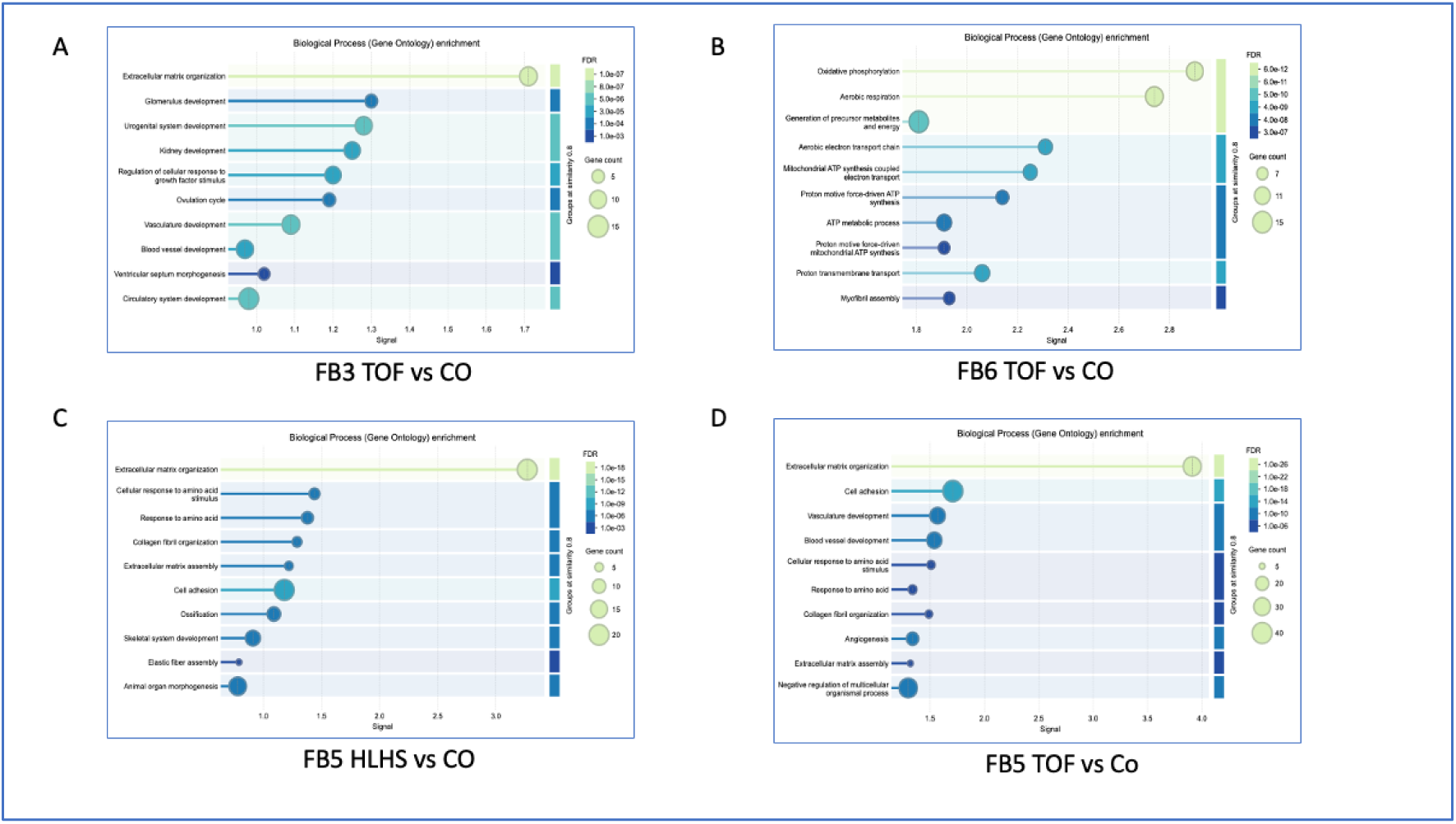
Gene Ontology enrichment analysis of the significantly de-regulated genes in cardiac fibroblasts (FBs). A. FB3 in tetralogy of Fallot (TOF) vs. controls (CO). B. FB6 in TOF vs. CO. C. FB5 in hypoplastic left heart syndrome (HLHS) vs. CO. D. FB5 in TOF vs. CO.

**Supplementary Figure S7.**
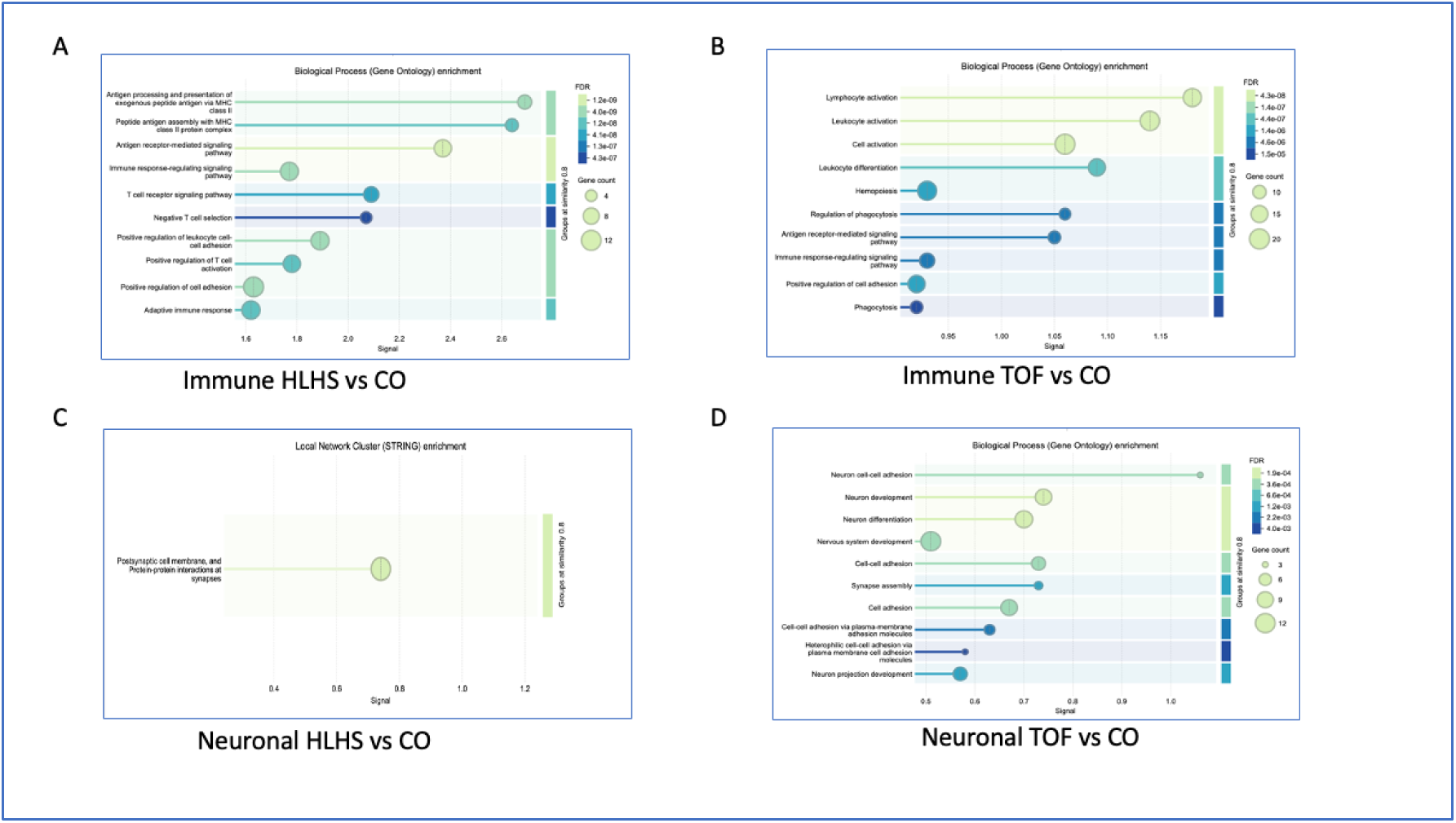
Gene Ontology enrichment analysis of the significantly de-regulated genes in immune and neuronal cells. A Immune, hypoplastic left heart syndrome (HLHS) vs. controls (CO). B. Immune, tetralogy of Fallot (TOF) vs. CO. C Neuronal, HLHS vs. CO. D Neuronal, TOF vs. CO.

**Supplementary Figure S8.**
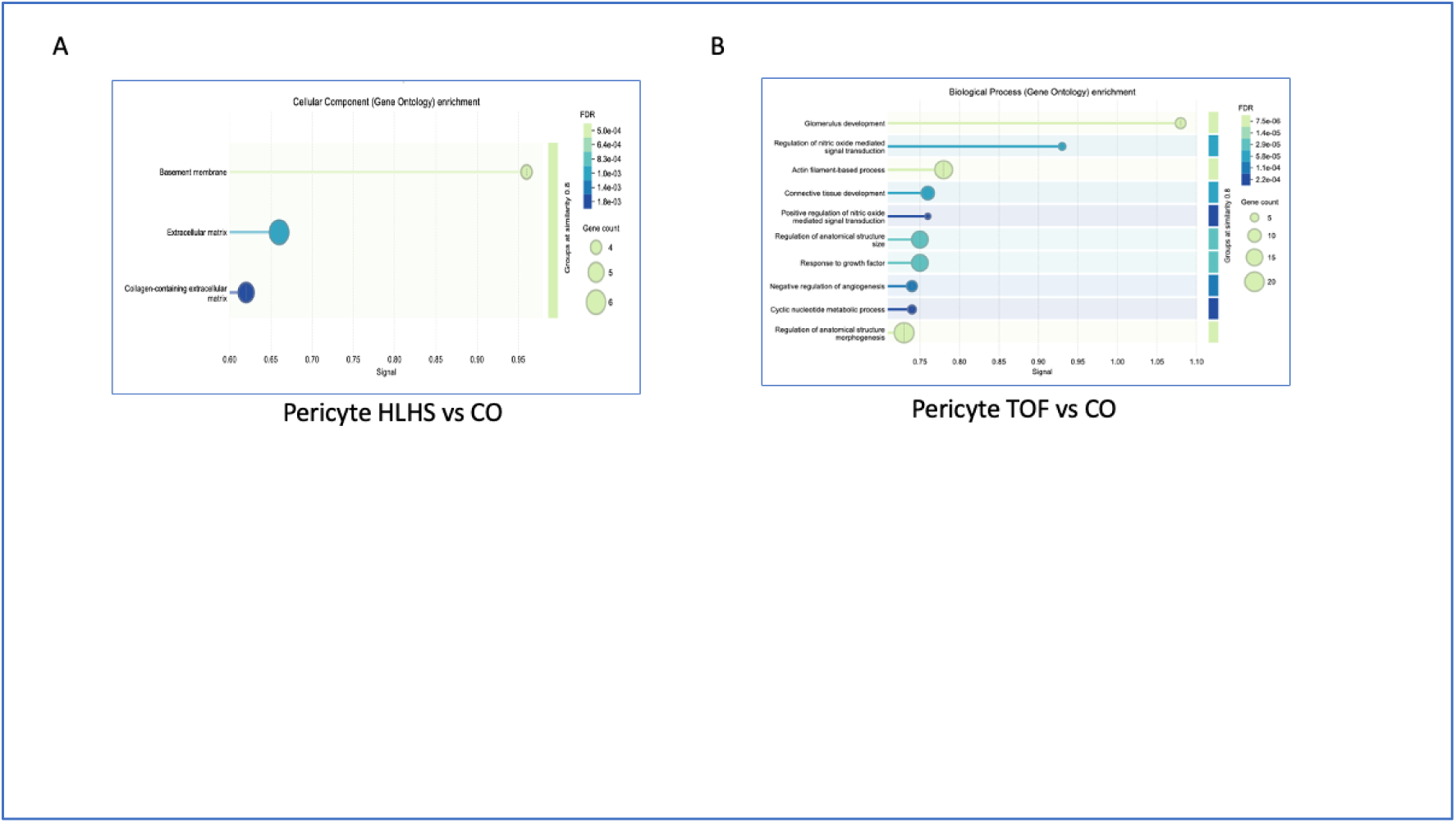
Gene Ontology enrichment analysis of the significantly de-regulated genes in pericyte subtypes in the pediatric cardiac biopsy samples. A. Hypoplastic left heart syndrome (HLHS) vs. controls (CO). B. Tetralogy of Fallot (TOF) vs. CO.

